# A within-host birth–death and time–dose–response model for Legionnaires’ disease

**DOI:** 10.1101/2025.01.31.25321481

**Authors:** Nyall Jamieson, Christiana Charalambous, David M. Schultz, Ian Hall

## Abstract

Understanding the dose–response relationship for infectious diseases is important for quantitative microbial risk assessment studies to mitigate risk. To capture the dose–response dynamics, understanding the pathogenesis of the infectious agent is desirable. Typically, attempting to understand the dose–response dynamics would involve within-host mathematical modelling and fitting dose–response curves to experimental data. No mathematical model exists that describes the within-host dynamics that occur within an individual infected with Legionnaires’ disease. Further, most dose–response models are based either on a single-hit or threshold hypothesis for the cause of illness. Here, we derive a model to explain within-host dynamics post-infection with Legionnaires’ disease that incorporates heterogeneity at the cellular and population levels. We develop a new dose–response model that allows for either of two hypotheses for the cause of illness, adding a new level of flexibility not currently seen in the literature. We extend the dose–response model to incorporate time as we develop a dose-dependent incubation-period model that is based on biological mechanisms. Our within-host models provide an ID50 of between eight and nine *Legionella* and median incubation periods close to four days, which is consistent with evidence obtained from animal experiments and human outbreaks in the literature.

## 1 Introduction

*Legionella* is a gram-negative bacterium that causes legionellosis [1]. Specifically, this disease may occur in both pneumonic (Legionnaires’ disease) and non-pneumonic (Pontiac disease) forms [2]. *Legionella* typically grows in freshwater or within water systems and, upon being aerosolised, can be inhaled and infect humans through the lungs [1]. After entering the lungs and inducing Legionnaires’ disease, the bacterium causes a range of symptoms such as fever, coughing, breathing difficulties, headache, nausea and death [1]. Additionally, unlike many other infectious diseases, Legionnaires’ disease is not believed to be a person-to-person transmissible disease [3]; instead, infection occurs exclusively through inhalation of aerosolized *Legionella* from contaminated water, which then settles in the lungs [3].

Like other intracellular pathogens such as *Coxiella burnetii* (the causative agent for Q-Fever) and *Francisella tularensis* (the causative agent for tularemia), *Legionella* reproduces inside host cells, rather than freely in the environment. Specifically, phagocytic cells within the human lungs attempt to eliminate *Legionella* through a process called *phagocytosis*. To survive, *Legionella* employ defence mechanisms that allow it to reside within phagocytic cells, where it reproduces until the host cell ruptures. Once the cell ruptures, *Legionella* are released and can infect additional cells. Each time this process occurs, proinflammatory cytokines in the lungs activate, recruiting more phagocytes as part of the adaptive immune response. This inflammatory response may trigger an onset of symptoms in an individual after a period of time (i.e., the incubation period).

Estimates of the incubation period have been determined in the literature, typically ranging from two to ten days [4, 5, 6, 7, 8, 9, 10]. Alternatively, a median incubation period of five days [11] or seven days [12] has been reported, with a mean incubation period of 5.3 days estimated when accounting for censoring issues associated with incubation-period data [13]. However, none of these approaches account for the dose of *Legionella* received, which may affect the incubation period. While no human dose-response data exists for Legionnaires’ disease, an experimental study in guinea pigs has provided insight into this relationship [14]. Guinea pigs exhibit similar pathological development and symptoms to humans [15, 16], and this similarity has made them a valuable resource for studying Legionnaires’ disease in humans. As a result, this experimental study [14] has formed the basis for quantitative microbial risk assessment studies, which estimate that between 11 and 12 *Legionella* are required to cause illness in 50% of individuals (ID50) [18, 17].

Mathematical models have been developed for various pathogens to better understand the relationship between dose received and probability of illness (dose–response models) [19, 20]. For these models, exact disease-specific mechanistic modelling is desirable, but these require comprehensive knowledge of the biological processes. Therefore, in many circumstances an exact model is not feasible to obtain. A typical approach to reduce the complexity is to assume that the infection process may be described using one of two common hypotheses. A single-hit hypothesis would assume that all inhaled *Legionella* act independently and are capable of causing a hit (i.e., after infection, each *Legionella* can independently trigger symptoms within the individual). Alternatively, a threshold hypothesis [19] would assume that an individual has a *threshold* of *Legionella* that can be initially deposited within their body without succumbing to illness and above which, illness occurs [20]. The choice of assumed hypothesis may significantly impact the results of fitting dose–response models to data, specifically for low-dose risk calculations [21]. Evidence exists to indicate that the single-hit hypothesis produces results consistent with observed infection DR data, specifically in the case of bacterial and viral agents [21]. Research has been conducted to extend DR models for Legionnaires’ disease to quantify the probability of illness based on both the dose received and the time elapsed since infection (known as time–dose–response models). However, the models that incorporate a time dependency are based on data from experimental studies, as opposed to being developed from any biological modelling [24, 22, 23].

No mathematical model currently describes the time–dose–response (TDR) relationship for Legionnaires’ disease in a mechanistic manner, nor does a mathematical model exist that describes the within-host dynamics of the infection process of Legionnaires’ disease. A model describing the entire infection process would require an understanding of both the innate and adaptive immune response to Legionnaires’ disease. The *Legionella pneumophila* pathogenesis, in the context of the host immune response, has been described providing an account of both the innate and adaptive immune response [25]. However, a mathematical model of the adaptive immune response would require accounting for the roles of cytokines. Developing such a model would likely include considerable levels of noise and highly correlated parameters that cannot be estimated from the data. Instead, we may ignore the role of cytokines, and model the number of extracellular *Legionella* as a surrogate for symptoms. This modelling strategy follows the approach used to develop within-host models of *Francisella tularensis* [26] and *Coxiella burnetii* [27], in which the simple birth–death model is extended to model intracellular bacterial growth within alveolar macrophages.

In this paper, we adapt the birth–death models developed for *Francisella tularensis* [26] and *Coxiella burnetii* [27] to develop a within-host model for Legionnaires’ disease. We then extend these models to account for stochasticity in the number of intracellular *Legionella* released upon macrophage death, as well as heterogeneity at both the cellular level and the required level of inflammation for symptom onset across the population. Our extended model will provide insight into the DR relation and the dose-dependent incubation period of Legionnaires’ disease. Additionally, we develop a new versatile DR model capable of accommodating either of the single-hit or threshold hypotheses. We incorporate time into this DR model based on valid biological mechanisms [13], enabling us to develop a mechanistic TDR model for Legionnaires’ disease. To our knowledge, this paper provides the first mathematical within-host model of Legionnaires disease, as well as the first such flexible DR model in the literature. Finally, we will compare our TDR model with the data-driven TDR counterpart [28, 24, 22, 23] to determine which approach provides a more preferable description of the TDR relation for Legionnaires’ disease.

## 2 Methods

In this section, we describe the assumptions and models used for developing both a within-host model of Legionnaires’ disease infection, as well as a DR and TDR model for Legionnaires’ disease. First, we describe the internal processes that occur once *Legionella* infects an individual and the simplifying assumptions that are made to model this biological process. Second, we develop a continuous-time Markov chain (CTMC) model alongside an analogous deterministic model to represent the within-host dynamics post-infection with *Legionella*, which is to be parameterised in the Results section. We describe the within-host models developed for *Francisella tularensis* [26] and *Coxiella burnetii* [27] and discuss our approaches for extending these models to account for stochasticity at the rupture level, as well as heterogeneity at the cellular and population level. We also provide details on the discrete-event simulation of these within-host models. Third, we introduce an intuitive framework for DR modelling, as we summarise common DR models. We then develop a versatile biologically-motivated DR model that bridges the gap between both hypotheses within the framework. Fourth, we extend the new DR model to incorporate time as we derive a biologically-motivated TDR model. We discuss alternative TDR models from the literature, which are based on experimental results or heuristic arguments, for comparison with our biologically-motivated TDR model.

### 2.1 Within-host dynamics of *Legionella* infection

In this paper, we consider short-term exposure to aerosols containing *Legionella*. An exposed individual may inhale aerosols that carry varying numbers of *Legionella*. These aerosols will travel to the lungs, where some of the *Legionella* may survive and establish themselves. Subsequently, phagocytic cells, such as alveolar macrophages, attempt to consume the *Legionella* through phagocytosis in order to cure the individual from their infection. As the *Legionella* and macrophages interact, cytokines are activated that recruit other phagocytes (e.g., neutrophils, monocytes, dendritic cells) to the lung as part of the adaptive immune response. These additional phagocytes contribute to the killing of *Legionella*. However, alveolar macrophages are the primary phagocytic cells involved during the *Legionella* infection process [25] and as such are considered as the sole phagocytic cell as we build the within-host model. As we only consider macrophages, we are only required to ascertain estimates for one type of phagocyte.

As phagocytosis occurs, the macrophage may successfully eliminate the *Legionella* or the *Legionella* may survive and live intracellularly within the macrophage. In the latter scenario, intracellular *Legionella* begin to replicate. The intracellular population increases until reaching a point at which the macrophage ruptures, releasing the *Legionella* back into the lungs (Fig. 1).

**Figure 1:**
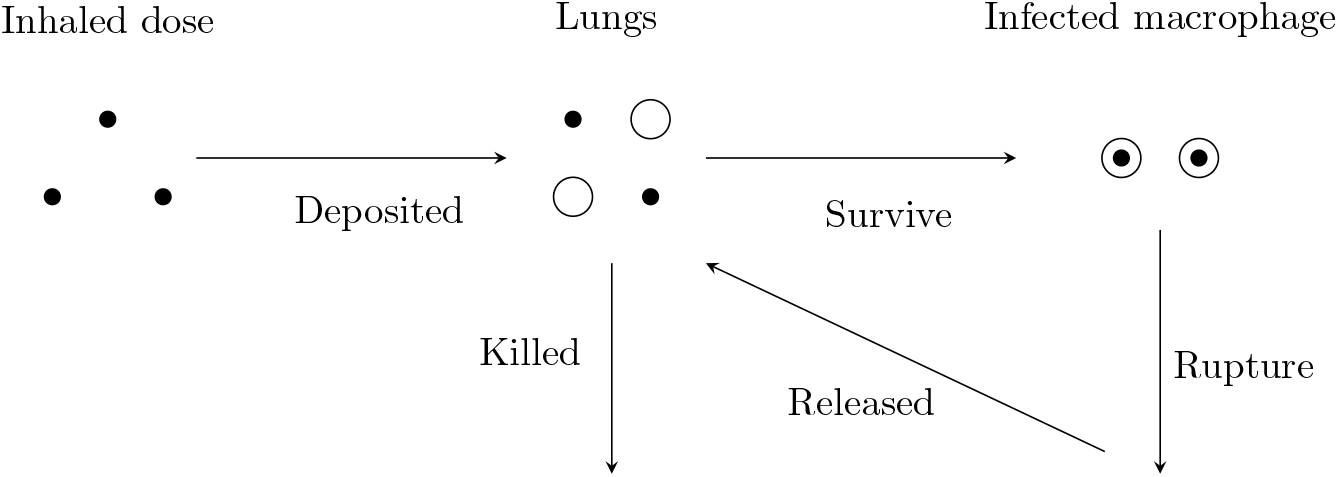
An illustration of the modelled dynamics post exposure to *Legionella*. Small black circles represent extracellular *Legionella* and large white circles represent macrophages. An individual is exposed to a dose of *Legionella*, some of which are inhaled. Inhaled *Legionella* may be deposited, successfully making their way to the lung. Of these *Legionella*, some may survive phagocytosis and infect a macrophage. After a certain period of time, the infected phagocytes may rupture. In this scenario, intracellular *Legionella* release back in the lungs.

The mechanisms that govern the number of viable macrophages in the lungs are unknown. Additionally, the number of alveolar macrophages within both lungs has been estimated to be of the order of 10^9^ [29]. Given the estimated large quantity of macrophages, we assume an infinite number of macrophages within the lungs, allowing for a parsimonious approach in which we develop a linear model of the infection process. This assumption simplifies the analysis by treating the macrophage population as approximately constant throughout the infection period. Nevertheless, in the electronic supplementary material, we relax the assumption and develop a more complex within-host model that accounts for a finite number of uninfected macrophages. In doing so, we derive a nonlinear model to describe the infection process, which converges to the linear model developed in Section 2 when the non-infected macrophage population is sufficiently large. With the estimated number of macrophages in the human lungs, the linear model presented in Section 2 remains appropriate for reasonably modeling the infection dynamics of Legionnaires’ disease.

We assume that any macrophage engulfs no more than one *Legionella* at a time. Therefore, a macrophage will either focus on killing the now-intracellular *Legionella* or will wait to rupture after engulfing the extracellular *Legionella*. Given the abundance of macrophages relative to extracellular *Legionella*, competition for engulfment likely prevents a single macrophage from engulfing more than one bacterium before others do so.

The exact mechanisms that results in macrophage rupture remains unclear; however, macrophages typically do not undergo apoptosis until later stages of infection (18-hours post infection) [30]. We assume a distribution for the mortality time of infected macrophages. Further, we model the intracellular *Legionella* population within an infected macrophage as a random variable with a mean dictated by a logistic growth process over time. This model reflects the early exponential growth within macrophages, dampened later by nutrient depletion due to increased intracellular *Legionella* competition for resources.

In reality, symptom onset results from heightened inflammation levels that are triggered by pro-inflammatory cytokines in response to phagocytosis and rupture events. For model parsimony, we use the extracellular *Legionella* population as a surrogate for the inflammation levels. We assume that a threshold *T*_*L*_ of extracellular *Legionella* within the lung is required for symptom onset.

### 2.2 Continuous-time Markov chain (CTMC) model

We introduce a CTMC approach for describing the process once *Legionella* deposit to the lungs. Following the within-host models for Francisella tularensis [26] and Coxiella burnetii [27], we consider a two-dimensional Markov chain (*L*(*t*), *M* (*t*)), with *L*(*t*) and *M* (*t*) representing the number of extracellular *Legionella* and infected macrophages respectively residing within the lungs at time *t* after infection. Three key events are modelled within the lungs: a macrophage killing an extracellular *Legionella* by phagocytosis, an extracellular *Legionella* surviving phagocytosis and infecting a macrophage, and an infected macrophage rupturing and releasing intracellular *Legionella* back in the lungs. The CTMC has two absorbing states: the individual is cured [(*L*(*t*), *M* (*t*)) = (0, 0)] or the individual has onset of symptoms [*L*(*t*) = *T*_*L*_].

Define *α* to be the rate at which an individual extracellular *Legionella* is engulfed and survives phagocytosis. Further, define *β* to be the rate at which an individual extracellular *Legionella* is engulfed and dies during phagocytosis. Therefore, *α* + *β* is the total rate of phagocytosis of an individual extracellular *Legionella* within the lungs. Additionally, define *λ* to be the rate at which an individual infected macrophage containing *Legionella* ruptures, releasing *Legionella* back into the lungs. These three events are modelled as Markovian random variables for model parsimony to fit within a CTMC framework.

Finally, we consider two scenarios for the rupture size (i.e., the number of *Legionella* released in a rupture event). First, we consider that *G Legionella* are released in a rupture event, which was the approach taken in [26, 27]. Second, the number of *Legionella* released in a rupture event follows the negative binomial distribution with mean *G* and dispersion parameter *θ*, which is an extension to the models developed in [26, 27]. In the former scenario, a state-change diagram is provided in [26, 27]. However, in the latter scenario, we visualise the state changes that may occur in the electronic supplementary material. The rates for state changes in the latter scenario are provided as follows:

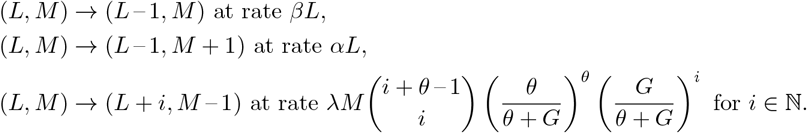

Alternatively, we replace the final rate with *λM* when it is assumed that *G Legionella* are released in every rupture event [26, 27]. Next, *d*_*I*_ is defined as the inhaled dose, and *ϕ* is defined as the probability that an individual inhaled *Legionella* is successfully deposited to the lung.

The Gillespie algorithm [31] is used to conduct the discrete-event simulation of the within-host model for Legionnaires’ disease until either absorbing state is reached. The simulation is varied for a deposited dose *d*_*D*_ ∈ (1, 500), where *d*_*D*_ ∼ Binom(*n* = *d*_*I*_, *p* = *ϕ*). We record 1000 iterations for each *d*_*D*_ to generate a probability of illness. Code that was used to run this simulation in R is provided in the electronic supplementary material.

A potential limitation of this CTMC is assuming that *α* and *β* are constant and independent of time. If we wish to consider the adaptive immune response and allow for other phagocytes to be recruited to the lungs, we may define *α* and *β* as functions of time. This change may likely result in the ratio *α/*(*α* + *β*) varying over time. In this case, a monotonically increasing function of time appears reasonable to incorporate the adaptive immune response. Parsimonious choices for such a function include a function proportional to the extracellular *Legionella* load, a function proportional to the number of infected macrophages, or a function proportional to the number of macrophage rupture events. However, due to our limited understanding of the effect that the adaptive immune response may have on a functional form, these options remain purely speculative. Moreover, there is insufficient data in the literature to parameterise any developed model of the adaptive immune response.

If we were to broaden the scope of our research to include multiple exposures of *Legionella*, we may extend the CTMC to allow for immigration of *Legionella* into the lungs post-infection. However, the timing of these immigration events depends solely on the individual’s exposure pattern and level of exposure. Furthermore, it is unclear whether the dynamics of multiple exposures can be described simply as an increase in extracellular *Legionella* populations with each additional dose, or if these dynamics are inherently more complex.

### 2.3 Deterministic model

When an infected macrophage ruptures, the extracellular *Legionella* population will increase by a large amount, the size of which varies between rupture events. Stochastic models are designed to capture these jumps and their variability, whereas a deterministic system only captures the average growth rate and fails to capture the jump in population size at low *Legionella* populations. If both the extracellular *Legionella* and infected macrophage populations increase, the stochastic system will have less variability and the rate of growth will tend towards that of a deterministic system. Therefore, a deterministic model is accurate for larger populations, but its inability to model the randomness means that a stochastic model must be used for smaller populations.

In spite of the shortcomings of a deterministic model, we develop a system of ordinary differential equations that describe the system in a deterministic manner for two reasons: the deterministic model provides a simple, computationally cheap method for describing population growth at large populations, and it allows us to obtain estimates for *α, β, λ* and *G* by fitting the differential equations to experimental data. The deterministic system of equations representing the populations *L* and *M* is given as follows:

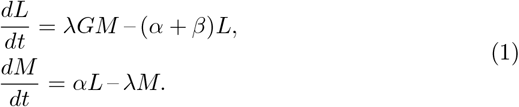

The solution to (1) with initial condition (*L*(0), *M* (0)) = (*ϕd*_*I*_, 0) is provided in (2), where 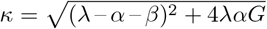 and 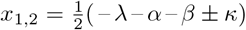. Because *G >* 0 then one of *x*_1,2_ must be negative and so we choose *x*_2_ *<* 0. Additionally, we require *x*_1_ *>* 0 to ensure that the extracellular *Legionella* population grows as the infection develops.

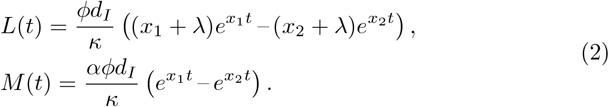

The second term on the right hand-side of the equation for *L*(*t*) in (2) decays to zero as *x*_2_ *<* 0. Because *x*_2_ *<* 0, rearranging (2) allows the approximate time at which illness occurs within the individual to be calculated [26], where *T*_*L*_ is the threshold number of extracellular *Legionella* required for onset of symptoms in the individual:

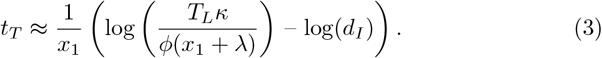

Further, (3) can be rearranged to consider a ‘low-dose’ incubation period (i.e. when *d*_*I*_ = 1), denoted *η*, as follows [26]:

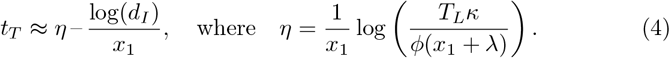

### 2.4 DR models

Mathematical DR models are defined as monotonically increasing, non-negative mathematical functions that calculate the probability of a response for a given dose, bounded by zero and one. These properties are satisfied by probabilistic cumulative distribution functions (c.d.f). Typically, mathematical DR models are fit to data from experimental studies to provide a functional form and further insight on the likelihood of illness. These experimental studies usually produce data that manifest as increasing when the proportion of individuals responding to a given dose is plotted against dosage [32]. Therefore, common probability distributions, often with sigmoidal shape, find usage for DR modelling [17, 19], although any c.d.f may serve this purpose. This sigmoidal relationship is analogous to the role of link functions in binomial regression, which connects predictors to the probability of illness in a non-linear manner. However, DR models should ideally be derived from biological mechanisms [21].

First, we begin by introducing a natural, easy-to-interpret framework for DR modelling. We provide an explanation of how common DR models fit within this framework. Second, we develop two new DR models, unique in their versatility to describe both the single-hit and threshold hypotheses. We apply these models to DR data generated from the discrete-event simulation of the within-host model to infer key properties of the DR curve for our model of Legionnaires’ disease. For example, we obtain the ID50 and find hypotheses for the DR relationship for Legionnaires’ disease.

In our research, we define a response as the onset of symptoms within an individual. Next, we define the illness parameter *z* of an individual, which has a different definition based on whether the single-hit or threshold hypothesis is assumed. In the single-hit scenario, *z* is defined as the probability that a single deposited *Legionella* results in onset of symptoms within the host. Alternatively, in the threshold scenario where the *Legionella* co-operates, a minimum threshold *z* ∈ (0, ∞) of *Legionella* is required for onset of symptoms within the host. For a deposited dose *j* ≥ *z*, illness occurs; for a dose *j < z*, the individual clears the infection.

Following this, we define *k*(*j*|*z*) as the probability that an individual, with illness parameter *z*, develops symptoms after exposure to a dose *j* [19]. Next, *q*(*z*) is the distribution of *z* which may model heterogeneity at the cellular or population level. With these distributions we develop a mixture distribution, the probability *r*(*j*) that an individual develops symptoms after subjected to a dose *j* is calculated as follows [19]:

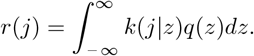

Here the distribution of illness parameters are assumed continuous but we assume dose is discrete. The function *g*(*j*|*d*_*I*_) is defined as the probability mass function (p.m.f) determining the dose *j* that an individual receives given the dose *d*_*I*_ that they are expected to receive [19]. The probability of symptom onset given an expected dose *P* (*d*_*I*_) is derived as follows:

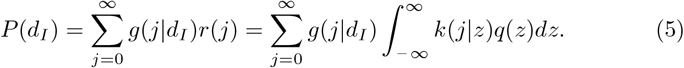

Next, we discuss modelling choices for *k*(*j*|*z*), *q*(*z*) and *g*(*j*|*d*_*I*_). Two main approaches may be used to derive a model for *k*(*j*|*z*). The choice of approach depends on which hypothesis is assumed for the mechanisms that result in an onset of symptoms from the infectious agent. In the single-hit scenario, if we assume a deposited dose *j* and probability *z* ∈ (0, 1), the term *k*(*j*|*z*) is derived by calculating the probability of at least one success from binomial distribution with *j* number of trials and *z* probability of success in each trial:

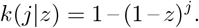

In the threshold scenario, the term *k*(*j*|*z*), as defined in [19], is given by a Heaviside function [33]:

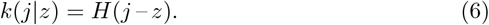

Next, *q*(*z*) can be given as a point mass fixed at *z* indicating that the illness parameter is fixed from host-to-host and bacteria-to-bacteria. For the single-hit model, a natural approach to model heterogeneity in the illness parameter is to assume that *z* ∼ Beta(*a, b*) to allow for a varying probability between *Legionella* or between hosts. Further, for the threshold hypothesis, *z* may be modelled with any continuous distribution *q*(*z*) valid over (0, ∞). In this case *r*(*j*) = *Q*(*j*), with *Q*(*z*) defined as the c.d.f corresponding to *q*(*z*). Therefore, *z* may be modelled with common population heterogeneity distributions (e.g., gamma, normal, log-normal, Weibull and Burr [19, 32, 34, 35, 36, 37, 38]).

Finally, various approaches to model the term *g*(*j*|*d*_*I*_) exist. One may assume that the inhaled dose equals the expected dose. In this scenario, a point-mass distribution, 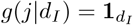 (*j*), is used. To allow for variability between the dose ingested and expected dose ingested, discrete count distributions may be used. In this scenario, we may use a Poisson distribution with mean *d*_*I*_, or a negative binomial distribution with mean *d*_*I*_ and overdispersion parameter *δ*:

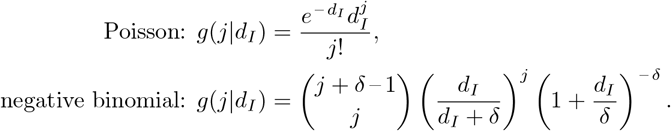

The choice between the Poisson and the negative binomial distribution is irrelevant for the single-hit model, which can be seen by highlighting that (5) may be rearranged as follows:

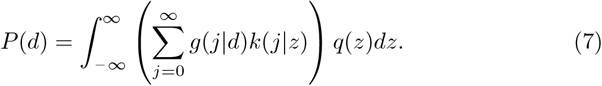

In the scenario of *g*(*t*|*d*_*I*_) ∼ Pois(*d*_*I*_), the sum within the bracket of (7) equals 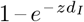 (the exponential DR model). In the scenario of *g*(*j*|*d*_*I*_) ∼ NB(*d*_*I*_, *δ*), the sum in (7) has the same functional form as the Poisson model with *z* replaced by *ζ* = *δ* log(1 + *z/δ*) [39]. Therefore, only the super-parameter *ζ* can be estimated, as opposed to the specific parameters making up *ζ*. Hence, both the Poisson or negative binomial models provide the same results, although *z* in the Poisson model and *ζ* in the negative binomial model offer different interpretations [39].

Three DR models are widely used in the literature: the exponential DR model, the beta-Poisson DR model (often confused with approximate Beta-Poisson model) and the Hill DR model [17, 20, 21, 22, 23, 24, 26, 27, 47, 40, 41, 42, 43, 44, 45, 46, 48]. Table 1 lists these DR models, with the various assumptions for *g*(*t*|*d*_*I*_), *k*(*j*|*z*) and *q*(*z*) required to derive each of them. These DR models will be used in analysis of the DR generated from the simulation of the within-host models.

**Table 1:** DR models obtained in the literature. The parameter space and definitions for *g*(*j*|*d*), *k*(*j*|*z*) and *f* (*z*) are given for clarity. We use *d* as opposed to *d*_*I*_ in this table for notational brevity. Here *d*_50_ is the parameter that describes the ID50 of the model. Additionally, the subscripts *b* and *h* in the dose–response *α* and *β* parameters represent the beta-Poisson and Hill dose–response models to distinguish these parameters with the *α* and *β* parameters from the within-host models.

| Model | Parameters | $P(d)$ | $g(j\vert d)$ | $k(j\vert z)$ | $q(z)$ |
| --- | --- | --- | --- | --- | --- |
| Exponential | $d_{50} > 0$ | $1 - e^{-d \log(2)/d_{50}}$ | $\text{Pois}(d)$ | $1 - (1 - z)^j$ | $\mathbb{I}_{[z=\kappa]}$ |
| Beta-Poisson | $\alpha_b, \beta_b > 0$ | $1 - {}_1F_1(\alpha_b, \alpha_b + \beta_b, -d)$ | $\text{Pois}(d)$ | $1 - (1 - z)^j$ | Beta |
| Hill | $\alpha_h, d_{50} > 0$ | $1/(1 + (\frac{d_{50}}{d})^{\alpha_h})$ | $\mathbb{I}_{[j=d]}$ | $H(j - z)$ | Log-logistic |

Next, we derive two DR models based on mechanistic justifications. After a single *Legionella* survives phagocytosis, the infected macrophage will eventually rupture. This rupture likely results in a large increase in the bacterial population. In the event where the initial dose is large and a rupture increases the population further, it is unlikely that all these surviving *Legionella* will be killed and extinction occurs. Therefore, for large doses, we expect that a single-hit model will be valid as one *Legionella* survival may be likely to eventually result in onset of symptoms.

However, this argument is less likely to be valid for smaller initial doses if *G* is not sufficiently large (with a probability (*β/*(*α* + *β*))^*G*^ of extinction of the released population following a rupture of size *G*). For small initial doses, a rupture may be unlikely to result in a *Legionella* population large enough for the overall probability of extinction to be nearly zero. As such, there is less justification to assume a single-hit description in this scenario. Thus, we consider the threshold hypothesis a viable alternative in this scenario. Further, we assume heterogeneity in the threshold across the population. Here we take the threshold distribution to be log-logistic as it is a common distribution for variability [45]. In summary, we assume that a single-hit model accurately describes the DR relationship for large initial doses, whereas a threshold model may more accurately describe the DR relationship for small initial doses.

To develop a DR model that does not assume an illness hypothesis a priori, we turn our attention to the Burr family of distributions for a DR model. The Burr family of distributions is defined as follows [13]:

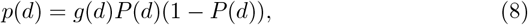

where *p*(*d*) is the p.m.f of the distributions, *P* (*d*) is the corresponding c.d.f for the distribution and *g*(*d*) is a function of the deposited dose. Consider a threshold *d*^*^ in which for low doses *d < d*^*^ we require a function that can reduce to the Hill DR model (i.e., *p*(*d*) = *α*_*B*_*P* (*d*)(1 – *P* (*d*))*/d*). Further, for large doses, *d > d*^*^, we require a function that tends to the exponential DR model (i.e., *p*(*d*) = *β*_*B*_(1 – *P* (*d*))). Such a DR model is obtained by assuming that *g*(*d*) = *α*_*B*_*/d* + *β*_*B*_ in (8). This model results in the functional form of the derived Burr distribution [13], but in the context of DR as opposed to incubation periods. We call this the Burr 1 DR model, defined as follows:

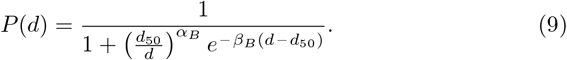

For the Burr 1 DR model, both *α*_*B*_ and *β*_*B*_ have an effect on the hypothesis for the DR phenomena. Estimates of *α*_*B*_ ≈ 0 indicate that the threshold hypothesis does not have validity in the given infection scenario. In this scenario, the model tends to the exponential DR model as *P* (*d*) → 1. Estimates of *β*_*B*_ ≈ 0 indicate that the single-hit hypothesis does not have validity in the given scenario. In this scenario, the model becomes the Hill DR model. Estimates of *α*_*B*_, *β*_*B*_≉; 0 indicate that both a single-hit and threshold model have some validity in the given scenario, depending on the initial dose deposited. Finally, *d*_50_ is the ID50.

Following this, we develop our second new DR model as we extend the Burr 1 DR model to allow for further flexibility. We first note that for the Burr 1 DR model, the Hill DR model is a special case as *β*_*B*_ = 0. However, although Burr 1 tends towards the exponential for large doses and *α*_*D*_ = 0, the exponential DR is not a special case. We extend Burr 1 by defining 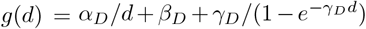 to develop the Burr 2 DR model. If *α*_*D*_ = *β*_*D*_ = 0, the third term in *g*(*d*) causes the Burr 2 to reduce exactly to the exponential DR model. Therefore, Burr 2 extends Burr 1 to include the exponential and Hill DR models as special cases. This choice of *g*(*d*) results in the following c.d.f for Burr 2:

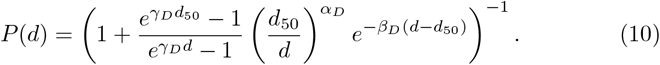

When fitting DR models to DR data simulated from the within-host models, we fit the three commonly used DR models defined in Table 1, as well as our two new Burr DR models.

### 2.5 Incorporating time into DR models

We begin this section by introducing TDR models and discussing some common approaches for TDR modelling in the literature. We then develop our own TDR model based on biological mechanisms.

Time–dose–response models are an extension of dose–response models that quantify the probability of a response (a response is defined as onset of symptoms) based on both the dose received and time elapsed since infection. A common approach for other diseases to incorporate time into a DR model, discussed in [49], is to include an incubation-period c.d.f *F* (*t*) in the exponent of the exponential DR model as *P* (*t, d*) = 1 – *e* ^−*zdF* (*t*)^. Here, *P* (*t, d*) is the probability of a response at a time *t* after exposure to a dose *d*. Based on heuristic arguments, *F* (*t*) may be modelled by the gamma, log-normal or Weibull distributions, as these are common incubation-period distributions. Alternatively, the Burr types III, X, XII or the derived Burr [13] incubation-period distributions offer alternatives built up from a mechanistic understanding of bacterial (or viral) growth dynamics [13]. Although *F* (*t*) ∼ Burr is derived from biological mechanisms [13], the implementation of a time dependency into the DR model in this way is based on mathematical convenience as opposed to biological validity. On the other hand, one distribution for *F* (*t*) may be derived from using a competing risk framework [49]. With this approach, one may assume that both the time until *Legionella* cause illness and the time until the *Legionella* become extinct are exponentially distributed. In this case, *F* (*t*) follows the exponential distribution [49]. This method provides a natural approach to incorporate time into the exponential DR model in a justifiable manner.

Additionally, one approach has been taken for analysis on Legionnaires’ disease experimental data that incorporates time into the exponential and beta-Poisson models [24, 23, 22, 28]. These models incorporate a time dependency in a data-driven way, as opposed to driven by any biological mechanisms (Table 2).

**Table 2:**
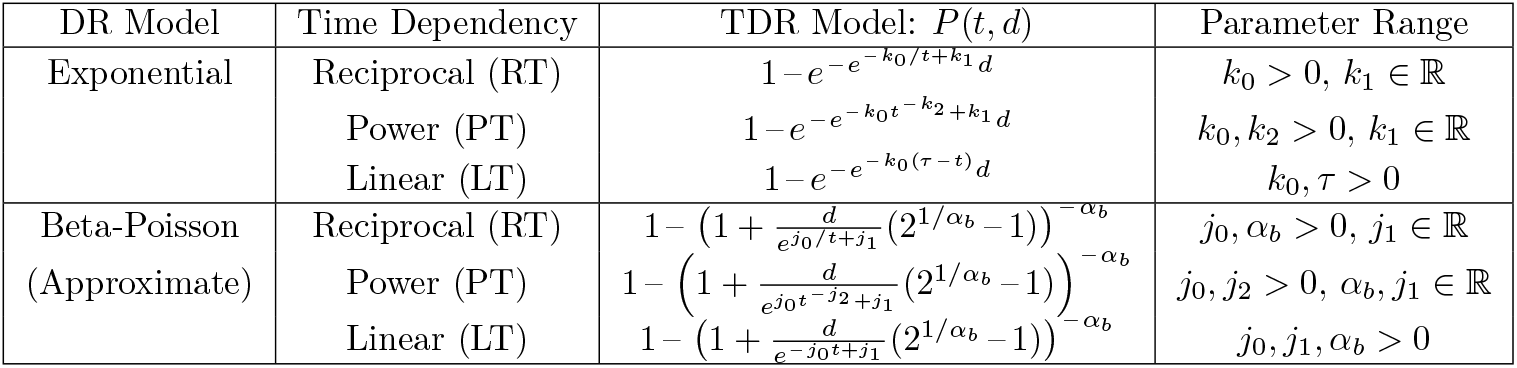
TDR models that have been developed for *Legionella* in the literature [24, 23, 22]. For *τ* in the exponential DR model with the LT time dependency, we note that although *τ >* 0, *τ* will be estimated to be sufficiently large in the fitting procedure so that *P* (*t, d*) ≈ 0 at *t* = 0.

We consider one final method for developing a TDR model. We write the law of conditional probabilities as follows:

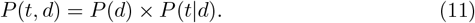

We model *P* (*d*) using the Burr 2 DR model and *P* (*t*|*d*), which is the dose-dependent incubation period, using the derived Burr incubation-period model [13] with a dose-dependent median incubation period. The Burr distribution for *P* (*t*|*d*) provides a biologically-valid dose-dependent incubation period that includes a natural way to define the median incubation period as a function of dose. The median incubation period is defined as a combination 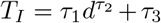 where *τ*_1_, *τ*_2_ and *τ*_3_ are model parameters.

We will fit the data-driven TDR models defined in Table 2, as well as our mechanistic conditional Burr TDR model defined in (11) to the data simulated from the within-host models for two reasons. First, we aim to gain insights into the dose-dependent incubation period of Legionnaires’ disease. Second, we aim to assess the validity of our mechanistic TDR response model compared to the data-driven TDR models developed in the literature [24, 23, 22].

## 3 Results

Now that we have derived the within-host model for Legionnaires’ disease, the Burr DR models, and the conditional TDR model, the next step is to run a simulation of the within-host model. We begin by obtaining estimates for the parameters in the within-host model. We make three choices in our approach of running the within-host model to compare different model assumptions. First, we consider a model (model A) in which we assume homogeneity at both the population and cellular level, but allow for stochasticity in the timing of the in-host events. In this scenario, the parameters used are the point estimates obtained (Table 3), which was the approach taken in [26, 27]. Second, we consider a model (model B) in which we assume stochasticity in the timing of the in-host events, as well as homogeneity as above for *λ, α*, and *β*. However, we allow for variability in the rupture size, in which we use the negative binomial distribution described earlier, which is an extension of the approach taken in [26, 27]. Third, we consider a model (model C) in which we assume a completely heterogeneous population of *Legionella*, macrophages and humans, which is an extension of the approach taken in [26, 27]. The host heterogeneity is incorporated by sampling *λ, G, α, β* and *T*_*L*_ from the appropriate distributions generated (Table 3), accounting for the dependencies between parameters (the method of which is discussed in the electronic supplementary material).

**Table 3:** A summary of the estimates obtained for the parameters in the deterministic extended birth–death model and the CTMC simulation.

| Event | Parameter | Units | Estimate | 95% CI |
| --- | --- | --- | --- | --- |
| Macrophage rupturing | $\lambda$ | 1/hours | 0.041 | (0.039, 0.042) |
| Macrophage carrying capacity | $\Psi$ | bacteria | 177.953 | (159.958, 195.903) |
| Intracellular *Legionella* growth rate | $\omega$ | hours | 0.192 | (0.165, 0.219) |
| Mean rupture size | $G$ | bacteria | 62.326 | (38.618, 90.412) |
| *Legionella* surviving phagocytosis | $\alpha$ | 1/hours | 0.089 | (0.009, 0.242) |
| *Legionella* dying by phagocytosis | $\beta$ | 1/hours | 1.088 | (0.113, 2.563) |
| Probability of deposition | $\phi$ | probability | 0.249 | (0.219, 0.280) |
| Low-dose incubation period | $\eta$ | hours | 108.955 | (98.478, 159.209) |
| Extracellular *Legionella* threshold | $T_L$ | bacteria | 50661 | (13452, 122967) |

The simulations for each within-host model are completed in the same way. For each dose, 1000 iterations of the stochastic simulation are completed. The proportion of iterations in which the host was considered to have developed symptoms is recorded and used as a probability that, for the given dose, the individual has developed symptoms. Further, the times at which symptom onset occurs is reported, for completing TDR analysis.

The results from simulating our three within-host models are used to obtain DR and TDR data of Legionnaires’ disease. In this section, we fit common DR models and our Burr DR model to the simulated DR data for Legionnaires’ disease. Similarly, we compare the results from fitting the data-driven TDR models with our conditional TDR model to the dose-dependent incubation-period data obtained from running the within-host simulation. In this section, we attempt to assess the DR relationship and dose-dependent incubation period for Legionnaires’ disease.

### 3.1 Parameterisation

The within-host birth–death model is now parameterised, as the parameters *λ, G, α, β, ϕ* and *η* are estimated from data in the literature. We consider two approaches for estimating parameters in a model: point estimates and distributional estimates. Point estimates for parameters *λ, G, α* and *β* are estimated from fitting to relevant datasets via a non-linear least squares approach, whereas a point estimate for *ϕ* is obtained from simulating *Legionella* deposition using the multiple-path particle dosimetry model (MPPD) software [50] and a point estimate for *η* obtained from fitting an accelerated failure time (AFT) model to interval-censored incubation-period data. Further, distributional estimates for each parameter are obtained through a bootstrapping approach and an assumption of normality in parameter estimates. Careful consideration is required to account for the dependencies of parameters, with a parameter dependency network provided in the electronic supplementary material.

#### 3.1.1 Rupture rate *λ*

We now proceed to the estimation of the within-host model parameters, beginning with *λ*, the reciprocal of which is defined as the median rupture time of infected macrophages. While the exact biological cause of macrophage rupture is unclear [25], we rely on data-driven approaches to estimate *λ* due to the lack of a clear mechanistic understanding. Specifically, *λ* is estimated using experimental data from a study in which bone marrow-derived macrophages from mice infected with wild type *Legionella* [51]. The percentage of ruptured macrophages was recorded hourly over a period of three days. This dataset informs our estimates of *λ*, as we fit common delay distributions to this dataset to model the observed mortality patterns.

We fit the exponential, gamma, log-normal, Weibull, Burr types III, X and XII, as well as the derived Burr distribution [13], to the experimental data of macrophage rupture times [51], with the derived Burr resulting in the best model fit (Fig. 2 (a)). Fitting the model and defining 1*/λ* as the median time for macrophage rupture, we obtain a point estimate of *λ* = 0.041 per hour. To obtain a distribution for this parameter, we bootstrap a dataset of size 1000 from the fitted Burr model and calculate *λ* as the reciprocal of the median of this dataset. We repeat 10000 times for a distribution of *λ*. We obtain a 95% bootstrap-generated confidence interval of (0.039, 0.042) for *λ*.

**Figure 2:**
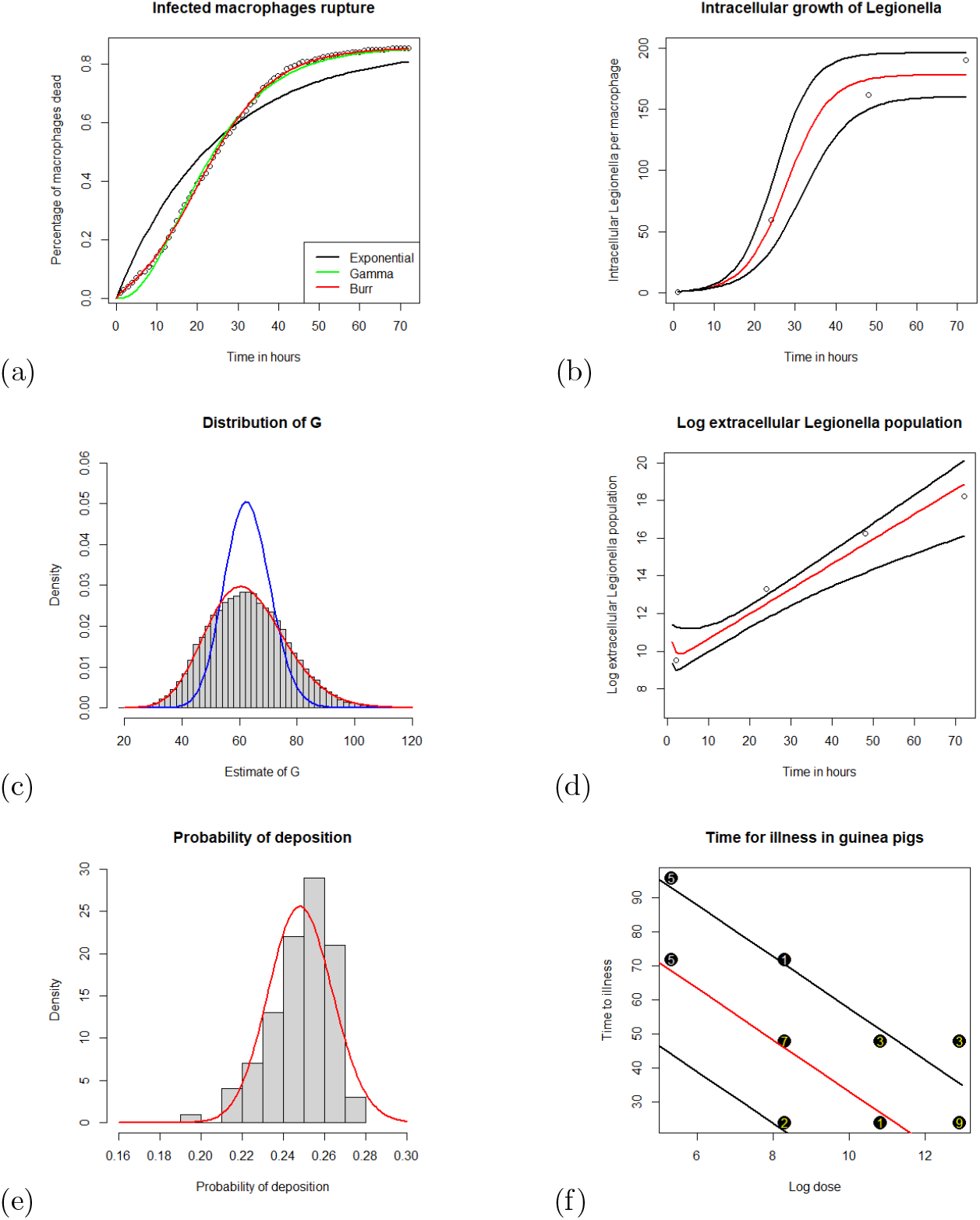
The sub-figure (a) represents the percentage of macrophages killed by *Legionella* hourly [51], with the derived Burr distribution fitted for estimation of *λ*. The sub-figure (b) represents the mean number of *Legionella* per macrophage after surviving phagocytosis and proliferating, with (12) fitted for estimation of Ψ, *ω* and *G*. The sub-figure (c) represents the rupture size distribution of a macrophage. The Poisson (*G* = 62.795 bacteria) and negative binomial distributions (*G* = 62.792 bacteria and *θ* = 32.072) are fit to the distribution of *G*, with blue and red curves respectively. The sub-figure (d) represents the number of *Legionella* within the lungs of mice [53], with (2) fitted for estimation of *λ, G, α* and *β*. The sub-figure (e) represents a histogram of the deposition results from inputting the parameter estimates into the MPPD software. A beta-distributed model fit to the MPPD data in red to estimate *ϕ*. The sub-figure (f) represents the time for illness in guinea pigs for varying doses, with (4) fitted data [55] to estimate *η*.

#### 3.1.2 Macrophage carrying capacity Ψ, intracellular growth rate of *Legionella ω* and mean number of *Legionella* released per rupture event *G*

Next, we estimate the number *G* of *Legionella* that are released back into the lungs following a rupture event. To do this, we use data from [52], in which U937-derived human macrophages were infected with *Legionella*. In this experiment, at 1, 24, 48 and 72 hours post-infection, the system was washed to remove extracellular *Legionella*, and the number of intracellular *Legionella* with the macrophages were recorded. Three repeated experiments yielded an average number of intracellular *Legionella* per macrophage at each time point.

To model the intracellular growth of *Legionella*, we use a piecewise logistic function [26, 27], which captures three distinct stages of bacterial replication within macrophages:

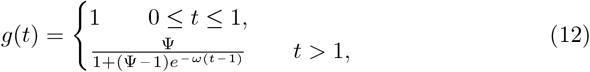

where Ψ represents the carrying capacity (the maximum number of *Legionella* within a macrophage) and *ω* represents the intracellular *Legionella* growth rate. The model assumes that for the first hour post-infection, the intracellular *Legionella* population remains constant at one, representing a delay before replication begins as the bacteria adapt within the macrophage and form a *Legionella*-specific phagosome. After this adaptation period, the bacteria enters a growth phrase, during which replication follows a logistic growth pattern. This phase is followed by a stationary phase, where competition and nutrition depletion within the macrophage cause the growth rate to slow.

We fit the logistic-growth model (12) to the intracellular *Legionella* data [52] using a nonlinear least squares approach to obtain point estimates for Ψ and *ω* (Fig. 2 (b)). Assuming normality in the non-linear least squares estimates we calculate distributions for these parameters. This model yielded an estimate of Ψ = 177.953 bacteria with a 95% confidence interval of (159.958, 195.903), as well as *ω* = 0.192 per hour with a 95% confidence interval of (0.165, 0.219). This estimate of *ω* indicates that the intracellular *Legionella* doubles every 3.6 hours during the logistic growth phase.

By combining our results for Ψ, *ω* and *λ*, we calculate the average number of *Legionella* released upon macrophage death as *G* = *g*(1*/λ*) = 62.326 bacteria. We calculate the corresponding *G* for each *λ*, Ψ and *ω* combination that was sampled from their distributions. This process generates a distribution of rupture sizes (Fig. 2 (c)). We obtain a 95% confidence interval of G as (38.618, 90.412). The Poisson distribution fails to accurately represent the variability observed in the rupture size distribution, as it cannot account for the overdispersion present in the data. Therefore, a negative binomial distribution provides a more suitable model fit (Fig. 2 (c)). In this case, we obtain a negative binomial with mean *G* = 62.792 bacteria and overdispersion *θ* = 32.072.

#### 3.1.3 Survival and mortality rates of *Legionella* in phagocytosis *α* and *β*

Furthermore, we consider an experiment [53] in which the number of *Legionella* within the lungs of mice was recorded at intervals 2, 24, 48 and 72 hours post-infection. Unlike other parameters, *α* and *β* are not deduced from the literature due to the large number of complex biological processes governed by innate and later adaptive immune responses. Instead, we estimate these parameters following the approach in [26]. Using the previously obtained estimates for *λ* and *G*, we fit the equations for the number of *Legionella* in (2) to the data using a non-linear least squares approach for estimating *α* and *β* (Fig. 2 (d)). To improve stability of the model-fitting procedure, we reparameterised (2) by introducing *γ* = *α* + *β* (the total rate of phagocytosis) and *π* = *α/*(*α* + *β*) (the probability that *Legionella* survives phagocytosis) and retransformed back to the *α, β* formulation once the model was fit. To obtain distributions for *α* and *β*, we sample from the corresponding distributions of *λ* and *G*, refit (2) using the sampled values of *λ* and *G*, and then assume normality in the resulting parameter estimates to derive distributions of *α* and *β*. This procedure yielded estimates of *α* = 0.089 per hour with a 95% confidence interval of (0.009, 0.242) and *β* = 1.088 per hour with a 95% confidence interval of (0.113, 2.563). We note a limitation in the dataset, which contains only 4 points. The fourth data point (at time 72 hours) may suggest that *Legionella* population growth may no longer be exponential by 72 hours. However, fitting a curve to the first three points (assuming exponential growth for the first 48 hours) still falls within a 95% confidence interval of the exponential-growth model that fits to all four points (Fig. 2 (d)).

#### 3.1.4 Probability of *Legionella* deposition *ϕ*

The next parameter that must be estimated, *ϕ*, is the probability of deposition into the human lungs. We take an approach similar to that of [27], where the MPPD software tool for estimating deposition [50] was used. For model parsimony, we assume an aerosol particle density of 1 gcm^−3^ and that the bacteria is spherical. *Legionella* bacteria has been reported to be of width *r*_1_ ∈ (0.5, 1)*µ*m and length *r*_2_ ∈ (1, 3)*µ*m [54]. We assume a spherical *Legionella* that has equal volume with radius *r*_3_. The diameter, *d*_3_ can be estimated as 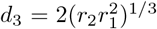. We use *r*_2_ = 1*µ*m as an upper bound of a confidence interval on the radii and *r* = 0.75*µ*m as the mean of the radii with a log-normal distribution, as it is stated to be the most common aerosol size distribution [27]. The particle size is modelled as log-normal with parameters *µ* = 0.732 and *σ* = 0.167.

Next, a model for the breathing rate of an individual must be used. Following [27], it is assumed that breathing rates are normally distributed with log-location equal to 0.012 and log-scale equal to 0.002. Estimates for the probability of deposition can be estimated by inputting these into the MPPD software. We then obtain 100 estimates, and a beta distribution is fitted to the data (Fig. 2 (e)), with parameter estimates given as *a* = 191.18 and *b* = 577.26. An estimate of *ϕ* = 0.249 is obtained to be the median of the resulting beta distribution. We obtain a 95% confidence interval of (0.219, 0.280).

#### 3.1.5 Low-dose incubation period *η* and extracellular *Legionella* illness threshold *T*_*L*_

After determining the values of *λ, G, α, β* and *ϕ*, we estimate the low-dose incubation period *η*, which is critical in estimating the extracellular *Legionella* threshold *T*_*L*_ that triggers symptom onset in an individual. We consider an experiment in which guinea pigs were infected with varying doses of *Legionella* [55]. This experiment provides empirical data on the time required for the onset of symptoms at different infection doses, which is crucial for estimating *η*. However, this dataset presents challenges that a nonlinear least squares fitting approach with (4) cannot handle. The incubation periods are interval censored, as the guinea pigs were assessed for symptoms at intervals of 24 hours (i.e., an incubation period of 72 hours indicates that illness occurred between 48 and 72 hours). To estimate *η*, we apply (4) and fit an AFT model to account for this interval censoring. With this AFT model, the choice of incubation-period distribution did not result in a statistically significant difference in estimates of *η*. This estimation process, with assuming a Gaussian-distributed incubation period, yields an estimate of *η* = 108.955 hours with a 95% confidence interval of (98.478, 159.209) (Fig. 2 (f)). The distributional estimate for *η* is obtained by making a normality assumption of parameter estimates in an AFT model, as well as resampling values of *λ, G, α* and *β* to generate samples of *x*_1_ to be used iteratively in the AFT model fitting procedure.

#### 3.1.6 Summary of model parametrization

With parameter estimates for *λ, G, α, β, ϕ* and *η*, we can derive an estimate for the median threshold of *Legionella* that cause illness in an individual. Substituting these estimates into (4) results in an estimate threshold of *T*_*L*_ = 50661 extracellular *Legionella* with a 95% confidence interval of (13452, 122967). The distribution for *T*_*L*_ is obtained by sampling from each of the parameter distributions that *T*_*L*_ depends on and accounting for parameter dependencies to propagate uncertainty. Using this threshold in (3) provides insight into the expected time until onset of symptoms for a given dose, as it provides a stopping point for a discrete-event simulation of the within-host dynamics until disease clearance or symptom onset. Table 3 lists the point estimates and confidence intervals obtained for each parameter.

Before conducting the within-host simulations and analysing the DR and TDR results, we provide a brief discussion of our approach for parameter estimation. Animal data were used at various points in the parameter estimation stage. First, data on mortality times of macrophages in mice were used to estimate *λ* [51]. Second, the extracellular *Legionella* populations within the lungs of mice [53] was used to estimate *α* and *β*. Third, time–dose–response data from guinea pig experiments [55] were used to estimate *η*. Animal data was used for two reasons. First, no human data exists in the same contexts that are required for estimating these parameters. Second, both guinea pigs and mice are understood to provide a reliable model for Legionnaires’ disease in humans. Guinea pigs have been considered as a model for Legionnaires’ disease [56] as there is similar pathological development and resulting symptoms in humans as in guinea pigs [15, 16]. Further, antigens have been found within guinea pig that are also present within humans post-infection with *Legionella* [57, 58]. Additionally, a strain of mice infected with *Legionella* suffer acute pneumonia within the first 48 hours post-infection, resembling human infection with Legionnaires’ disease [59, 60]. Because of these reasons, we assume that these parameter estimates are feasible to a human application but more experimental work is needed to corroborate the theoretical insights.

### 3.2 DR results from the within-host simulations

The DR models are fitted to the simulated data for all three within-host models, which were defined at the beginning of Section 3) (Table 4. For simulating the within-host model A, the single-hit DR models provide a better fit to the data than the threshold model, based on AIC (Table 4). Therefore, the single-hit hypothesis appears more reasonable than the threshold hypothesis to model the DR relationship for Legionnaires’ disease when assuming a deterministic rupture size and homogeneous population (model A). If one *Legionella* succeeds in a hit, then a median of 62 intracellular *Legionella* are released back into the lungs. Either the extracellular *Legionella* will be killed by macrophages or will survive phagocytosis and live intracellularly within its host. By using a competing risks framework with the point estimates for *α* and *β*, the probability that a *Legionella* survives phagocytosis, given that it has not yet been killed via phagocytosis, is given as *α/*(*α* + *β*) = 0.076.

**Table 4:** Results of fitting the DR models to the simulated data, with the residual sum of squares and parameter estimates provided. The standard errors of estimates are provided in the electronic supplementary material.

| DR model | Stochastic model |  |  |  |  |  |
| --- | --- | --- | --- | --- | --- | --- |
|  | A |  | B |  | C |  |
|  | AIC | Parameter estimates | AIC | Parameter estimates | AIC | Parameter estimates |
| Exponential | -4206.51 | $d_{50} = 8.84$ | -4354.27 | $d_{50} = 8.95$ | -4094.97 | $d_{50} = 8.97$ |
| Beta-Poisson | -4205.38 | $\alpha_b = 212.62$<br>$\beta_b = 2489.22$ | -4352.32 | $\alpha_b = 1190$<br>$\beta_b = 14166$ | -4212.32 | $\alpha_b = 17.61$<br>$\beta_b = 201.42$ |
| Hill | -2910.94 | $\alpha_h = 1.76$<br>$d_{50} = 8.54$ | -2960.37 | $\alpha_h = 1.77$<br>$d_{50} = 8.66$ | -2924.17 | $\alpha_h = 1.72$<br>$d_{50} = 8.52$ |
| Burr 1 | -4319.69 | $\alpha_B = 0.79$<br>$\beta_B = 0.06$<br>$d_{50} = 8.90$ | -4332.84 | $\alpha_B = 0.87$<br>$\beta_B = 0.06$<br>$d_{50} = 9.01$ | -4400.20 | $\alpha_B = 0.81$<br>$\beta_B = 0.06$<br>$d_{50} = 8.84$ |
| Burr 2 | -4331.41 | $\alpha_D = -0.14$<br>$\beta_D = 0.08$<br>$\gamma_D = -0.07$<br>$d_{50} = 8.861$ | -4386.67 | $\alpha_D = 0.04$<br>$\beta_D = 0.08$<br>$\gamma_D = -0.13$<br>$d_{50} = 8.89$ | -4398.20 | $\alpha_D = -0.19$<br>$\beta_D = 0.06$<br>$\gamma_D = -0.0001$<br>$d_{50} = 8.84$ |

Similarly, the probability that a *Legionella* is killed by a macrophage is *β/*(*α* + *β*) = 0.924. If just one *Legionella* survives phagocytosis, then 62 bacteria will be released back into the lungs. For the individual to be cured, all 62 *Legionella* must be killed. Extinction following a single hit occurs with probability at most (*β/*(*α* + *β*))^*G*^ = 0.007. Hence, the survival of one *Legionella* is *likely* to be sufficient to cause illness. However, with model A, there is still over a 0.7% chance that extinction later occurs after a single hit. Therefore, although a single-hit model offers a reasonable approximation of the DR relationship for Legionnaires’ disease, for lower doses the single-hit model is less likely to be applicable. Both Burr DR models outperform both single-hit models, which supports the fact that the single-hit hypothesis can be improved upon by considering a combination of both hypotheses (Table 4).

Similar conclusions are drawn from the results of analysing within-host model B (Table 4). With the rupture size following a negative binomial distribution, a probability of 0.012 that a single-hit does not result in illness is obtained. Therefore, the single-hit hypothesis provides a good description of the internal dynamics. However, this percentage is not negligible, meaning that a single hit does not guarantee onset of symptoms. Therefore, a combination of hypotheses appears beneficial, which is supported by the fact that the Burr 2 model provides preferable model fits, based on AIC (Table 4). Again, the Hill model does not provide a good fit to the data, indicating that a threshold hypothesis on its own does not hold for Legionnaires’ disease (Table 4).

Further, the results of simulating the within-host model C indicate that all DR models provide a worse fit to the simulated data, compared to fitting to within-host models A and B (Table 4). The heterogeneity introduced simulating by sampling parameter estimates from distributions before each iteration of the within-host simulation result in the tested DR models struggling (relative to fitting to within-host models A and B) to capture the DR relationship (Table 4). Again, the single-hit models provide a better description than the threshold model. However, Burr 1 provides a better fit than all commonly used DR models, regardless of the hypothesis for the cause of illness (Table 4). Additionally, for within-host model C, extending Burr 1 to Burr 2 does not offer further improvements in model fit, as Burr 2 reduces to Burr 1 for this dataset (Table 4).

The Burr 1 DR model offers mixed results compared to the currently used single-hit DR models (Table 4). For within-host models A and C, the Burr 1 outperforms the exponential and beta-Poisson DR models based on AIC. However, for within-host model B, the Burr 1 is outperformed by both the exponential and beta-Poisson DR models based on AIC (Table 4). On the other hand, we obtain less variable results for the more complex Burr 2 DR model (Table 4). For within-host models A, B and C, the Burr 2 outperforms all commonly used DR models based on AIC (Table 4). These results suggest that developing a flexible model that does not assume a hypothesis for the cause of illness a priori offers a preferable alternative.

To evaluate the validity of our within-host models, we compare our estimated Legionnaires’ disease DR models with DR curves from guinea pig experiments reported in the literature [14] to assess the consistency of our results with the current understanding of the DR of Legionnaires’ disease. In this experiment, guinea pigs were exposed to various doses of *Legionella* (one, five, fifty and one hundred organisms), and the proportion of guinea pigs that developed symptoms was recorded: zero out of four, one out of four, eight out of eight and eight out of eight, respectively, for each dosage. The experimentally obtained DR curve is plotted, alongside the DR data obtained from simulating all three of our within-host models (Fig. 3).

**Figure 3:**
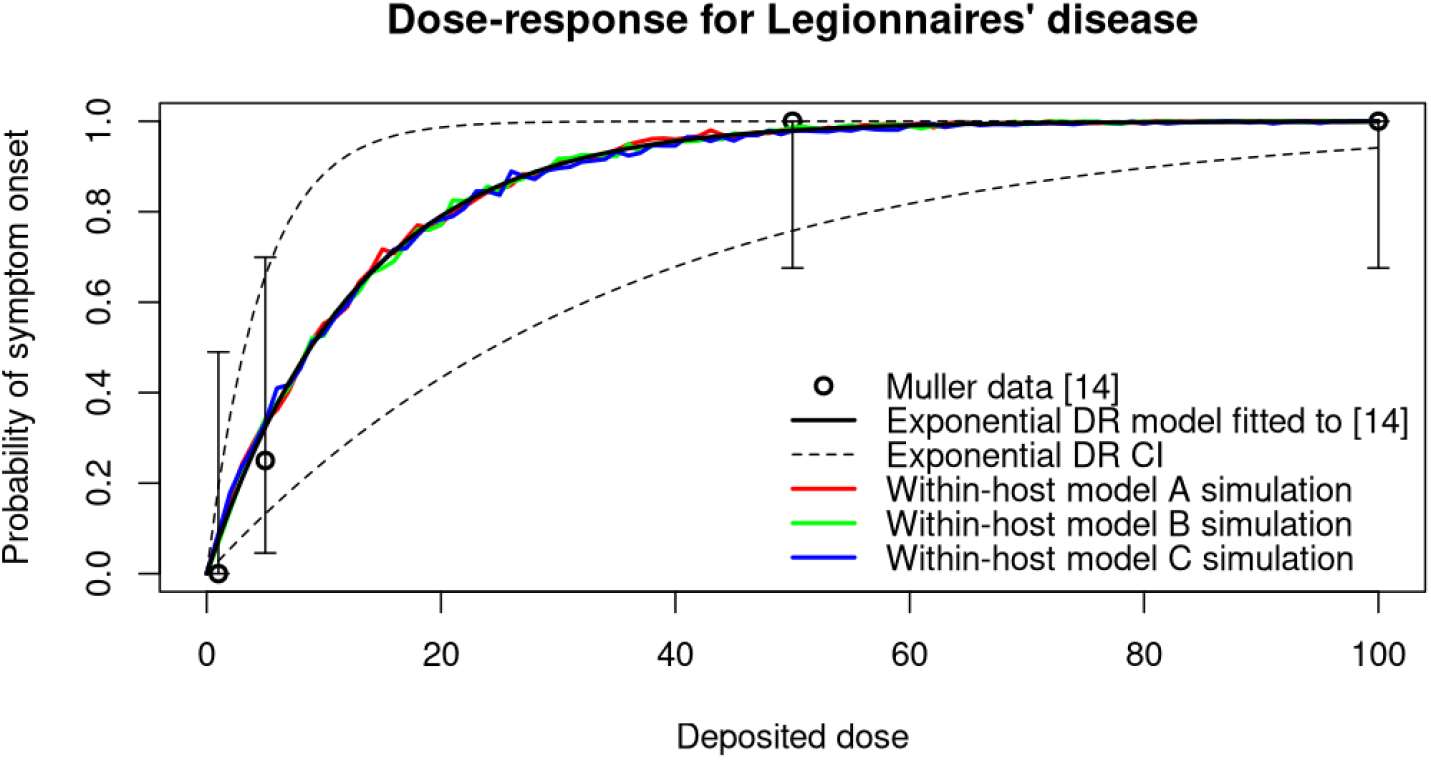
A plot of the guinea pig DR data as four data points representing the proportion of guinea pigs that developed symptoms [14]. Error bars are provided for each point from the [14] dataset to provide the uncertainty in their values when used to model the probability of onset of symptoms in this experiment. An exponential DR model is fitted to the data in [14] using a binomial likelihood approach. The DR data obtained from simulating the three within-host models are presented for comparison (red, green and blue respectively).

As the three within-host models of Legionnaires’ disease were developed independently of the experimental DR data [14], comparisons between the DR curves can be made without risk of a circular argument. All three within-host models provide DR results consistent with evidence observed in previous experiments (Fig. 3). All three within-host models predict an ID50 that falls within the 95% confidence interval of (3.22, 24.44) estimated from animal experiments, which was estimated by fitting an exponential dose–response model to the data using a binomial likelihood approach (Fig. 3) [14]. Specifically, model A predicts 8.86 with a 95% confidence interval of (8.80, 8.91), model B predicts 8.89 with a confidence interval of (8.84, 8.95) and model C predicts 10.00 with a 95% confidence interval of (9.23, 10.76) (Fig. 3). Moreover, the DR curves obtained from simulating the three within-host models lie within the 95% confidence interval from the exponential DR model fitted to [14] for the full range of doses, aligning closely with the estimated DR curve fitted to the experimental data [14]. These findings provide evidence supporting the validity of our within-host models and their ability to capture the DR dynamics of Legionnaires’ disease.

#### 3.2.1 TDR analysis

This section begins by providing the results from fitting the six previously developed TDR models for Legionnaires’ disease, as well as our newly-derived conditional probability Burr TDR model, to the data generated from simulating the three within-host models (Table 5). Among the data–driven models developed in [24, 23, 22], the power time (PT) dependency consistently provided the best results across all models, regardless of whether the exponential or approximate beta-Poisson DR model was applied (Table 5).

**Table 5:**
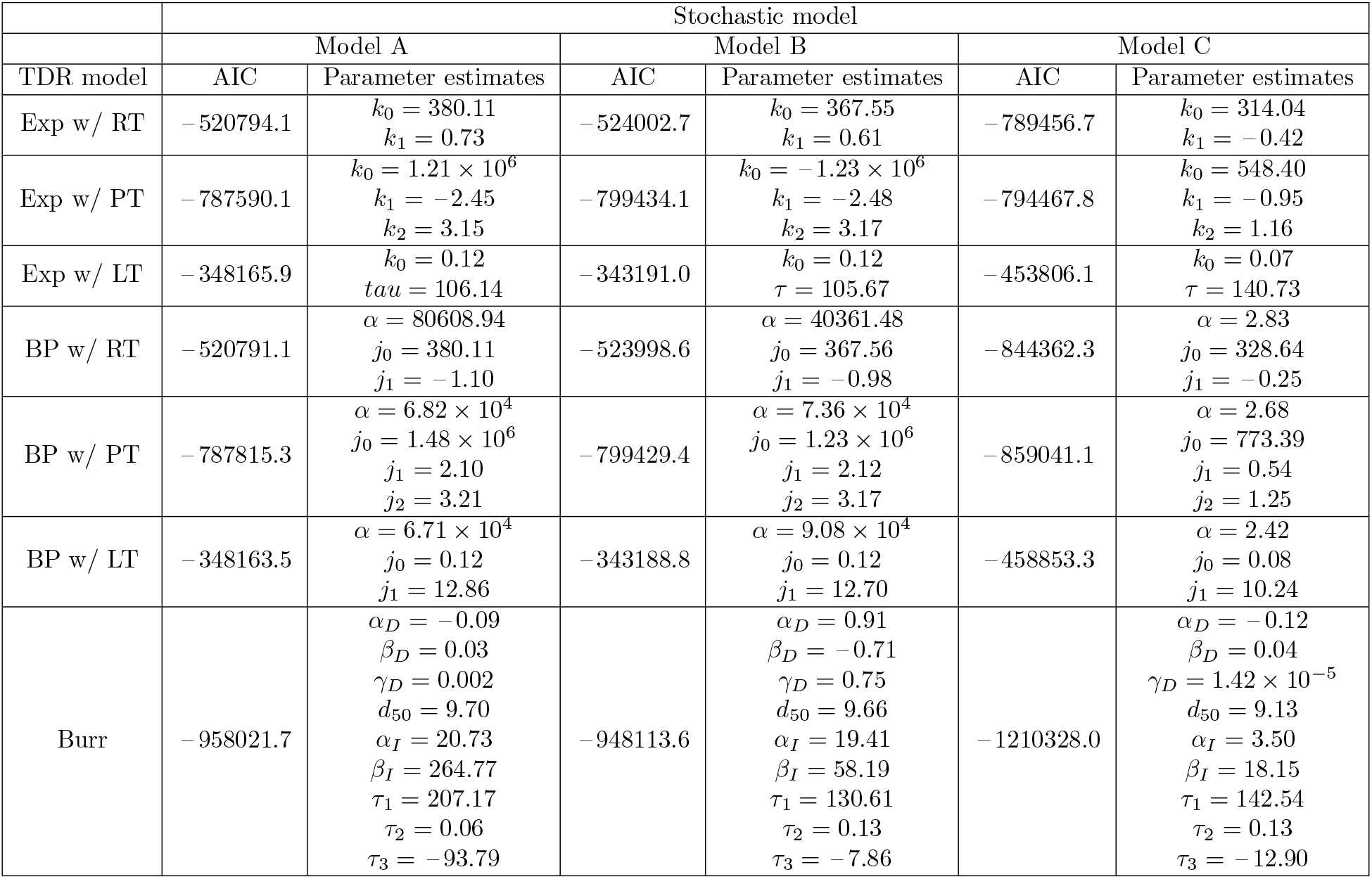
The results of fitting both the data–driven and mechanistic TDR models to the simulated data, with the AIC and parameter estimates provided. The standard errors of estimates are provided within the electronic supplementary material.

After comparing the results from fitting the six previously developed TDR models to the data simulated from the three within-host models, we now assess how these results compare with those obtained from our conditional Burr TDR model (Table 5). For all three within-host models, the conditional probability Burr TDR model outperforms all previously developed Legionnaires’ disease TDR models based on AIC (Table 5). The Burr 2 DR model, combined with the derived Burr incubation period—designed to make the median incubation period dose-dependent—provides improved results for TDR modeling of Legionnaires’ disease (Table 5). Additionally, as well as outperforming the data-driven models defined in Table 2, our Burr TDR model (11) provides a mechanistic derivation for a TDR model, which is based on biological mechanisms for both the dose–response component and dose-dependent incubation period component of the conditional probability model (11).

Next, we consider the dose-dependent incubation-period distributions that are obtained from simulating each of the three within-host models. A probability heat-map is provided for each within-host model (Fig. 4). For all three within-host models, a clear pattern exists with a decrease in the median incubation period as the deposited dose increases (Fig. 4). At the ID50, the mean incubation period appears to be close to four days for all within-host models (confidence intervals for the mean incubation period at the ID50 were obtained for within-host models A, B and C, yielding intervals of (3.78, 3.96), (3.83, 4.02) and (3.89, 4.19) days respectively. While all models effectively capture the median incubation period, the incubation period from within-host models A and B center heavily about the median for larger initial doses (Fig. 4). For low doses, models A and B somewhat capture the variability of incubation periods that has appeared in human incubation period data [4, 5, 6, 7, 8, 9, 10, 11, 12, 13]. Further, model C may offer a distribution that has a broader range and less pronounced peak at larger doses (Fig. 4), which more closely aligns with human outbreak data [4, 5, 6, 7, 8, 9, 10, 11, 12, 13].

**Figure 4:**
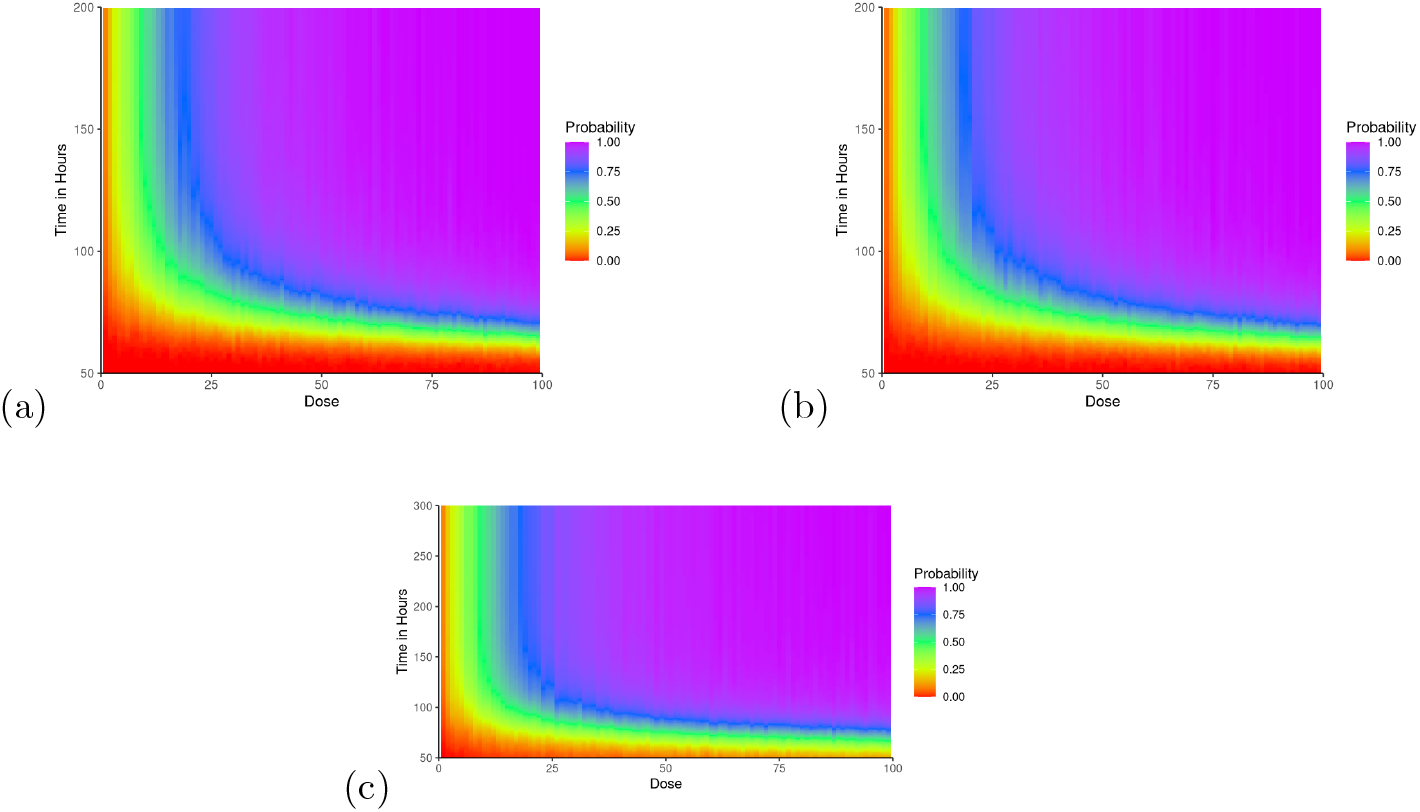
The TDR results obtained from simulating the three within-host models of Legionnaires’ disease. Sub-figure (a) represents within-host model A. Sub-figure (b) represents within-host model B. Sub-figure (c) represents within-host model C.

These results indicate a clear time dependency between the incubation period and the initial dose deposited for Legionnaires’ disease. To quantify this relationship, we compute Spearman’s correlation *τ*_*i*_ (for model *i* ∈ *{*A, B, C*}*) between the initial deposited dose and the time until symptom onset. The results reveal strong negative correlations: *τ*_*A*_ = – 0.65, *τ*_*B*_ = – 0.64 and *τ*_*C*_ = – 0.66, indicating that high deposited doses are associated with shorter incubation periods in all three within-host models. These findings align with biological expectations, as larger deposited doses likely result in more *Legionella* initially surviving phagocytosis and reproducing, leading to accelerated growth in the early post-infection period. However, a large amount of *Legionella* are required to survive phagocytosis and subsequently rupture a macrophage in order to reach *T*_*L*_ = 50661. Therefore, although the deposited dose affects the incubation period, the time required for all these phagocytosis and subsequent rupture events to occur plays plays an important role on the incubation period of Legionnaires’ disease. This is further illustrated in the sensitivity analysis provided within the electronic supplementary material.

## 4 Discussion

This paper introduces and develops models for three distinct issues with modelling the within-host dynamics post-infection with an infectious disease. First, we extended the model developed for *Francisella tularensis* [26] and *Coxiella burnetii* [27], in which we allow for heterogeneity at the cellular and population levels, as well as allow for a more realistic rupture size distribution of the infected macrophages, though we have had to use animal data to parameterize the model. We applied this to Legionnaires’ disease to gain an understanding of the DR and dose-dependent incubation period of this disease. Second, we have discussed common mathematical DR models that use the framework described in (5) as we have summarised the assumptions used in developing these models. We have developed two new biologically-motivated DR models that bridge the gap between both hypotheses for the cause of symptoms within an infected individual. We have fit the DR models developed in the literature, as well as our own, to data generated from simulating the within-host model. We use this to quantify the DR relationship for Legionnaires’ disease, as well as explore the validity of our Burr DR models. Third, we have developed a new TDR model based on conditional probabilities that allows for use of different DR and dose-dependent incubation-period models independently. We have compared this to the data-driven models developed for Legionnaires’ disease [22, 23, 24] to explore the validity of this conditional probability based approach.

Interestingly, our results offer insight into the DR hypothesis for Legionnaires’ disease. Notably, we found evidence to suggest that the single-hit hypothesis does not reliably describe the process for developing symptoms of Legionnaires’ disease. While an estimated median of 358 and 130 bacteria are released during macrophage rupture for the *Francisella tularensis* [26] and *Coxiella burnetii* [27] models respectively, the rupture size for *Legionella* is much lower. In other words, for these other diseases, the probability of symptom onset is essentially one after a rupture event. In contrast, for Legionnaires’ disease, a single hit does not guarantee symptom onset. Legionnaires’ disease appears unique in that both the single-hit and threshold hypotheses hold some validity. Therefore, neither the single-hit or threshold hypothesis reliably describing the process of developing symptoms of Legionnaires’ disease provides justification for the more flexible Burr DR models. Additionally, we applied the within-host models (A, B, and C) to predict the ID50 for symptom onset, revealing results consistent with experimental observations in the literature. Specifically, within-host models A, B, and C predict an ID50 of 8.86 (8.80, 8.91), 8.89 (8.84, 8.95), and 10.00 (9.23, 10.76) *Legionella* deposited in the lungs for symptom onset, respectively. These predictions align with results from DR experiments in the literature [14], where the estimated ID50 equals 8.87 *Legionella* with a 95% confidence interval of (3.22, 24.44).

Next, we explore the TDR results obtained in this research, as comparisons can be made with our TDR analysis and incubation periods from human outbreaks obtained in the literature. Our estimate for the low-dose incubation period is *η* = 108.955 hours, or 4.5 days. This estimate falls slightly below the lower end of the commonly reported mean of five to seven days, but still within the commonly stated incubation period of two to ten days [4, 5, 6, 7, 8, 9, 10, 11, 12]. Our results may highlight potential limitations in using guinea pig data to parameterise a human model. Perhaps using guinea pigs or mice data to infer parameter estimates for the Legionnaires’ disease infection process of humans is insufficient to describe individuals with immune system that falls short to that of the average human. Alternatively, the estimate of *η* may highlight bias and underestimation in incubation periods estimated from human outbreaks due to a double censoring issue of not having exact infection and symptom onset times [13].

Delving deeper into the incubation periods obtained from the within-host models, neither in-host randomness nor population heterogeneity account for the variability in incubation periods alone. Only by considering both sources of variability (within-host model C), we obtain an incubation-period distribution that corresponds more realistically to data obtained in the literature [4, 5, 6, 7, 8, 9, 10, 11, 12, 13]. Further, the results of our stochastic simulation indicate that for doses that typically cause illness, the mean time at which illness occurs appears to be closer to four days when considering any of the three within-host models. The incubation-period distributions obtained from simulating all three within-host models ranges from one to seventeen days, mainly falling within two to ten days. These results are consistent with human outbreaks with a two to ten day incubation period commonly reported [4, 5, 6, 7, 8, 9, 10, 11, 12], as well as the study of an outbreak in Melbourne in which incubation periods ranged from one to sixteen days [13]. Both the deterministic and stochastic models provide supporting evidence that the results of our research matches current beliefs in the literature.

Our analysis of the DR and TDR results from the within-host simulations aligns with current literature, suggesting that while incorporating additional complexities could enhance the model’s realism, such complexities are not essential for developing a reliable model of Legionnaires’ disease. To maintain model parsimony, we have made several simplifying assumptions regarding the dynamics post-infection. Specifically, we have excluded cytokine mechanisms involved in inflammation and used the extracellular *Legionella* population as a proxy for inflammation levels. Furthermore, we have not differentiated between various activation states of macrophages [61] and have assumed that all macrophages uniformly search for and phagocytose *Legionella*. Additionally, we have not accounted for other phagocytes or their recruitment to the lung due to uncertainties regarding the timing and magnitude of the adaptive immune response.

In the development of the within-host model, we have compared the differences between employing a deterministic model and stochastic model. The deterministic model results in exponential growth and inevitable symptom onset. However, unlike the stochastic model, the deterministic model does not allow for an individual to clear infection. Further, this model does not capture the variable rupture sizes, which has an effect on the population growth at small extracellular *Legionella* populations. As such, a stochastic discrete-event model is more favourable early after infection, whilst a deterministic model will be more suitable after a period of time has passed post-infection. The process will reach a point of no return, in which exponential growth approximately holds true and the individual will inevitably develop symptoms. Future research may quantify the time required post-infection for a deterministic to model the within-host dynamics reasonably well.

Focusing on the stochastic within-host model, we assumed exponentially distributed times for each event so that we can employ a Markovian framework. With the assumption that bacteria and macrophages move randomly within the alveolar region of the lungs, appealing to a Markovian distribution for phagocytosis events appears reasonable. However, other distributions provide a better fit to the macrophage rupture time data than the exponential distribution (Fig. 2 (a)). Therefore, although commonly chosen [27, 26], the exponential distribution is an oversimplistic modelling approach for macrophage rupture. To improve the model, other distributions may be applied for this event that would result in modelling with a non-Markovian framework. In this scenario, a zero-inflated model would be required as the probability of macrophage rupture does not tend to one. Alternatively, one may extend this model to use an Erlang distribution and use Erlang’s method of separating compartments to better describe the time to rupture [62, 63, 64].

In addition to the main findings, we discuss the assumptions that were made in estimating *ϕ*. We have assumed that each *Legionella* is inhaled separately when determining *ϕ*. In reality, multiple *Legionella* may be contained within an individual aerosol that is inhaled [1]. Increasing the aerosol size results in a different estimated inflammation threshold *T*_*L*_, as *T*_*L*_ ∝ *ϕ*. However, as *T*_*L*_ is in the order of 10^5^ and the number of extracellular *Legionella* required for symptom onset to be inevitable will be much lower than *T*_*L*_, the DR produced by a within-host model that assumes an increased aerosol size with varying number of *Legionella* would not differ from our results. Further, the time until symptom onset would likely vary, but not by a large amount. Because we expect a deterministic model to be valid before *T*_*L*_ is reached, we see that the expected increase in incubation period is proportional to log(*T*_*L*_*/T*_*ϕ*_) hours, where *T*_*L*_ is the estimate for our current model and *T*_*ϕ*_ is the extracellular *Legionella* threshold for a model in which *ϕ* is changed. This change will not have a large effect on the incubation period given that the median incubation period is estimated to be around four days.

In summary, this paper provides the first mathematical within-host model that describes the infection process of Legionnaires’ disease. We have obtained results for the DR curve and ID50, as well as low-dose incubation period, median incubation period and incubation period distribution (within-host model C) that agree with observed data in the literature. Additionally, we have developed two new DR models that provide more flexibility than currently used DR models. DR models in the literature assume a single-hit or threshold hypothesis a priori, whereas for the first time we develop a model that allows the hypothesis for the cause of symptoms to be inferred from the data. Moreover, we derive a mechanistic TDR model, based on combining our Burr DR model and the biologically justified Burr incubation period model. This approach allows us to model the TDR dynamics of Legionnaires’ disease and incorporate a dose-dependency within the incubation period. Our DR and TDR models offer improvements and more reliable understanding than by modelling with the data-driven models developed for Legionnaires’ disease within the literature.

## Data Availability

All data produced in the study are available at:
https://github.com/NyallJamieson/Within-host-model

## Acknowledgment

NJ acknowledges support from the Engineering and Physical Sciences Research Council (EPSRC) and Mathematics and Data in Scientific and Industrial Modelling (MADSIM) at the University of Manchester for funding of their studentship.

IH was supported by the JUNIPER modelling consortium (grant MR/V038613/1) the National Core Study on Transmission (PROTECT) and by the UKRI Impact Acceleration Account (IAA 386). NJ and IH also acknowledge the UK Health Security Agency (UKHSA) for honorary contracts and funding (for IH). The views expressed are those of the author(s) and not necessarily those of the Department of Health or UKHSA.

## References

[1] Prussin AJ, Schwake DO, Marr LC. Ten questions concerning the aerosolization and transmission of Legionella in the built environment. Building and Environment. 2017;123:684–695.

[2] World Health Organisation. Available from: https://www.who.int/news-room/fact-sheets/detail/legionellosis.

[3] Phin N, Parry-Ford F, Harrison T, Stagg HR, Zhang N, Kumar K, Lortholary O, Zulma A, Abubakar I. Epidemiology and clinical management of Legionnaires’ disease. The Lancet Infectious Diseases. 2014;14(10):1011–1021.

[4] Braeye T, Echahidi F, Meghraoui A, Laisnez V, Hens N. Short-term associations between Legionnaires’ disease incidence and meteorological variables in Belgium, 2011-2019. Epidemiology Infection. 2020 04;148:e150.

[5] De Giglio O, Fasano F, Diella G, Lopuzzo M, Napoli C, Apollonio F, Brigida S, Calia C, Campanale C, Marzella A, Pousis C, Rutigliano S, Triggiano F, Caggiano G, Montagna MT. Legionella and legionellosis in touristic-recreational facilities: Influence of climate factors and geostatistical analysis in Southern Italy (2001-2017). Environment Research. 2019 11;178:108721.

[6] Dunn CE, Rowlingson B, Bhopal RS, Diggle P. Meteorological conditions and incidence of Legionnaires’ disease in Glasgow, Scotland: application of statistical modelling. Epidemiology Infection. 2013 Apr;141(4):687–96.

[7] Fisman D, Lim S, Wellenius GA, Johnson C, Britz P, Gaskins M, Maher J, Mittleman MA, Victor Spain C, Haas CN, Newbern C. It’s not the heat, it’s the humidity: Wet weather increases legionellosis risk in the Greater Philadelphia Metropolitan Area. The Journal of infectious diseases.

[8] Gleason JA, Kratz NR, Greeley RD, Fagliano JA. Under the Weather: Legionellosis and Meteorological Factors. Ecohealth. 2016 06;13(2):293–302.

[9] Halsby KD, Joseph CA, Lee JV, Wilkinson P. The relationship between meteorological variables and sporadic cases of Legionnaires’ disease in residents of England and Wales. Epidemiology Infection. 2014 Nov;142(11):2352–9.

[10] Ricketts KD, Charlett A, Gelb D, Lane C, Lee JV, Joseph CA. Weather patterns and Legionnaires’ disease: a meteorological study. Epidemiology Infection. 2009 Jul;137(7):1003–12.

[11] Karagiannis I, Brandsema P, Van Der Sande M. Warm, wet weather associated with increased Legionnaires’ disease incidence in the Netherlands. Epidemiology Infection. 2009 Feb;137(2):181–7.

[12] Beauté J, Sandin S, Uldum SA, Rota MC, Brandsema P, Giesecke J, Sparen P. Short-term effects of atmospheric pressure, temperature, and rainfall on notification rate of community-acquired legionnaires’ disease in four European countries. Epidemiology and infection. 2016 Dec. Available from: https://pubmed.ncbi.nlm.nih.gov/27572105/.

[13] Jamieson N, Charalambous C, Schultz D M., Hall I. (2024). The Burr Distribution as a Model for the Delay between Key Events in an Individual’s Infection History. PLOS Computational Biology. 2024.

[14] Muller D, Edwards M L, Smith D W. Changes in iron and transferrin levels and body temperature in experimental airborne legionellosis. Journal of Infectious Diseases. 1983. doi:10.1093/infdis/147.2.302.

[15] Davis GS, Winn JR WC, Gump DW, Craighead JE, Beaty HN. Legionnaires’ pneumonia in guinea pigs and rats produced by Aerosol Exposure. Chest. 1983. doi:10.1378/chest.83.5.15s.

[16] Davis GS, Winn Jr WC, Gump DW, Beaty H. The kinetics of early inflammatory events during experimental pneumonia due to Legionella pneumophila in guinea pigs. Journal of Infectious Diseases. 1983. doi:10.1093/infdis/148.5.823.

[17] Armstrong TW, Haas CN. A quantitative microbial risk assessment model for legionnaires’ disease: Animal Model selection and dose-response modeling. Risk Analysis. 2007;27(6):1581–1596.

[18] Madera-García V, Mraz AL, López-Gálvez N, Weir MH, Werner J, Beamer PI, Verhougstraete MP. Legionella pneumophila as a health hazard to miners: A pilot study of water quality and qmra. Water, 11(8):1528, Jul 2019. doi: 10.3390/w11081528.

[19] Kodell RL, Kang SH, Chen JJ. Statistical models of health risk due to microbial contamination of foods. Environmental and Ecological Statistics. 2002;9(3):259–271.

[20] Teunis PF, Havelaar AH. The beta poisson dose-response model is not a single-hit model. Risk Analysis. 2000;20(4):513–520.

[21] Haas CN, Rose JB, Gerba CP. Wiley. Quantitative microbial risk assessment. 2014.

[22] Huang Y, Haas CN. Quantification of the relationship between bacterial kinetics and host response for monkeys exposed to aerosolizednbsp; nbsp; nbsp; nbsp; nbsp; nbsp; Francisella tularensis. Applied and Environmental Microbiology. 2011;77(2):485–490.

[23] Huang Y, Haas CN. Time-dose-response models for microbial risk assessment. Risk Analysis. 2009;29(5):648–661.

[24] Huang Y, Bartrand TA, Haas CN, Weir MH. Incorporating time postinoculation into a dose-response model ofyersinia pestisin mice. Journal of Applied Microbiology. 2009;107(3):727–735.

[25] Liu X, Shin S. Viewing Legionella pneumophila pathogenesis through an immunological lens. Journal of Molecular Biology. 2019;431(21):4321–4344.

[26] Wood RM, Egan JR, Hall IM. A dose and time response markov model for the in-host dynamics of infection with intracellular bacteria following inhalation: With application to Francisella tularensis. Journal of The Royal Society Interface. 2014;11(95):20140119.

[27] Heppell CW, Egan JR, Hall I. A human time dose response model for Q fever. Epidemics. 2017;21:30–38.

[28] Prasad B, Hamilton KA, Haas CN. Incorporating time-dose-response into Legionella outbreak models. Risk Analysis. 2016;37(2):291–304.

[29] Hume PS, Gibbings SL, Jakubzick CV, Tuder RM, Curran-Everett D, Henson PM, Smith BJ, Janssen WJ. Localization of macrophages in the human lung via design-based stereology. American Journal of Respiratory and Critical Care Medicine. 2020:1209–1217. 10.1164/rccm.201911-2105.

[30] Abu-Zant A, Santic M, Molmeret M, Jones S, Helbig J, Abu Kwaik Y. Incomplete activation of macrophage apoptosis during intracellular replication of Legionella pneumophila. Infection and immunity. 73(9), 5339–5349.

[31] Gillespie DT. A general method for numerically simulating the stochastic time evolution of coupled chemical reactions. Journal of Computational Physics. 1976;22(4):403–434.

[32] Gadagkar SR, Call GB. Computational tools for fitting the hill equation to dose–response curves. Clinical & Experimental Pharmacology. 2015;05(04).

[33] Weisstein, EW. Heaviside Step Function. Available at: https://mathworld.wolfram.com/HeavisideStepFunction.html.

[34] Turner ME. Some classes of hit-theory models. Mathematical Biosciences. 1975;23(3–4):219–235.

[35] Furumoto WA, Mickey R. A mathematical model for the infectivity-dilution curve of tobacco mosaic virus: Theoretical considerations. Virology. 1967;32(2):216–223.

[36] Pinsky PF. Assessment of risk from long term exposure to waterborne pathogens. Environmental and Ecological Statistics. 2000;7(2):155–175.

[37] Farber JM, Ross WH, Harwig J. Health risk assessment of listeria monocytogenes in Canada. International Journal of Food Microbiology. 1996;30(1–2):145–156.

[38] Burr IW. Cumulative frequency functions. The Annals of Mathematical Statistics. 1942;13(2):215–232.

[39] Pratt A, Bennett E, Gillard J, Leach S, Hall I. Dose–response modeling: Extrapolating from experimental data to real-world populations. Risk Analysis. 2020.

[40] Hill AV. A new mathematical treatment of changes of ionic concentration in muscle and nerve under the action of electric currents, with a theory as to their mode of excitation. The Journal of Physiology. 1910;40(3):190–224.

[41] Goutelle S, Maurin M, Rougier F, Barbaut X, Bourguignon L, Ducher M, Maire P. The hill equation: A review of its capabilities in pharmacological modelling. Fundamental amp. Clinical Pharmacology. 2008;22(6):633–648.

[42] Reeve R, Turner JR. Pharmacodynamic models: Parameterizing the hill equation, Michaelis-menten, the logistic curve, and relationships among these models. Journal of Biopharmaceutical Statistics. 2013;23(3):648–661.

[43] Seefeldt SS, Jensen JE, Fuerst EP. Log-logistic analysis of herbicide dose–response relationships. Weed Technology. 1995;9(2):218–227.

[44] Goldoni M, Johansson C. A mathematical approach to study combined effects of toxicants in vitro: Evaluation of the Bliss Independence Criterion and the Loewe additivity model. Toxicology in Vitro. 2007;21(5):759–769.

[45] Nussbeck FW. Log-Logistic models. Encyclopedia of Quality of Life and Well-Being Research. 2014:3686–3689.

[46] Goyal A, Reeves DB, Cardozo-Ojeda EF, Schiffer JT, Mayer BT. Viral load and contact heterogeneity predict SARS-COV-2 transmission and super-spreading events. eLife. 2021;10.

[47] Xu J, Carruthers J, Finnie T, Hall I. Simplified within host and DR models of SARS-COV-2. Journal of Theoretical Biology. 2022.

[48] Ke R, Zitzmann C, Ribeiro RM, Perelson AS. Kinetics of SARS-COV-2 infection in the human upper and lower respiratory tracts and their relationship with infectiousness. 2020.

[49] Brookmeyer R, Johnson E, Barry S. Modelling the incubation period of anthrax. Statistics in Medicine. 2005;24(4):531–542.

[50] Price O. MPPD: Multiple-path particle dosimetry model. 2023.

[51] Speir M, Vogrin A, Seidi A, Abraham G, Hunot S, Han Q, Dorn GW, Masters SL, Flavell RA, Vince JE, Nader J. Legionella pneumophila strain 130B evades macrophage cell death independent of the effector SIDF in the absence of Flagellin. Frontiers in Cellular and Infection Microbiology. 2017.

[52] Xiong L, Yamasaki S, Chen H, Shi L, Mo Z. Intracellular growth and morphological characteristics of lt;Igt;Legionella pneumophilalt;/igt; during invasion and proliferation in different cells. Biological & Pharmaceutical Bulletin. 2017;40(7):1035–1042.

[53] Gobin I, Susa M, Begic G, Hartland EL, Doric M. Experimental Legionella longbeachae infection in intratracheally inoculated mice. Journal of Medical Microbiology. 2009;58(6):723–730.

[54] Bouwknegt M, Schijven JF, Schalk JAC, de Roda Husman AM. Quantitative risk estimation for a Legionella pneumophila infection due to whirlpool use. Risk Analysis. 2012;33(7):1228–1236.

[55] Fitzgeorge RB, Baskerville A, Broster M, Hambleton P, Dennis PJ. Aerosol infection of animals with strains of Legionella pneumophila of different virulence: Comparison with intraperitoneal and intranasal routes of infection. Journal of Hygiene. 1983;90(1):81–89.

[56] Padilla-Carlin JD, McMurray DN, Hickey AJ. The Guinea Pig as a Model of Infectious Diseases. Comparative medicine. 2008.

[57] Maiwald M, Schill M Stochasinger, Helbig JH, Lück PC, Witzleb W, Sonntag HG. Detection ofLegionella DNA in human and guinea pig urine samples by the polymerase chain reaction. European Journal of Clinical Microbiology, Infectious Diseases. 1995. doi:10.1007/bf02112614.

[58] Williams A, Lever M. Characterisation of antigen in urine of guinea pigs and humans with legionnaires’ disease. Journal of Infection. 1995. doi:10.1016/s0163-4453(95)92691-7.

[59] Brown AS, van Driel IR, Hartland EL. Mouse models of legionnaires’ disease. Current Topics in Microbiology and Immunology. 2013.

[60] Brieland J, Freeman P, Kunkel R, Chrisp C, Hurley M, Fantone J, Engleberg C. Replicative Legionella pneumophila lung infection in intratracheally inoculated A/J mice. A murine model of human Legionnaires’ disease. The American Journal of Pathology. 1994.

[61] Hu G, Christman JW. Editorial: alveolar macrophages in lung inflammation and resolution. Frontiers in Immunology. 2019. doi:10.3389/fimmu.2019.02275.

[62] Bolzoni L, Della Marca R, Groppi M. On the optimal control of SIR model with Erlang-distributed infectious period: Isolation strategies. Journal of Mathematical Biology. 2021. 10.1007/s00285-021-01668-1.

[63] Sherborne N, Blyuss KB, Kiss IZ. Dynamics of multi-stage infections on networks. Bulletin of Mathematical Biology. 2015:1909–1933. 10.1007/s11538-015-0109-1.

[64] Cunniffe NJ, Stutt RO, van den Bosch F, Gilligan CA. Time-dependent infectivity and flexible latent and infectious periods in compartmental models of plant disease. Phytopathology. 2021:365–380. 10.1094/phyto-12-10-0338.

