## Appendix for "A within-host birth–death and time–dose–response model for Legionnaires’ disease"

### S1 Appendix

This appendix provides details of further analysis not included in the paper. First, we provide a derivation of the deterministic within-host model. Second, we provide a figure of how the within-host parameters depend on each other throughout the data-fitting procedure. Third, we provide plots of the distributions of the within-host parameters that are estimated from the experimental datasets. Fourth, we provide the standard errors that were obtained when fitting the dose–response and time–dose–response models to the data simulated from the within-host models. Fifth, we provide analysis on how the first four moments of the incubation-period datasets simulated from the three within-host models. Sixth, we provide the hierarchical analysis conducted on the incubation periods simulated from the three within-host models and the resulting clustered incubation-periods from each within-host model. Seventh, we provide results from the sensitivity analysis to investigate how varying the within-host model parameters affects the dose–response and time–dose–response results.

#### Derivation of a deterministic within-host model for Legionnaires' disease

In this appendix, we derive a deterministic model for the within-host dynamics that occur between infected macrophages and extracellular *Legionella* once an individual is infected with Legionnaires' disease.

Consider a biological system in which three populations exist: a population of uninfected macrophages $U$, a population of macrophages infected with *Legionella* $M$, and a population of extracellular *Legionella* $L$ that are free to move about the lungs. We define the total population at time $t$ as $N(t) = U(t) + M(t) + L(t)$. The term describing the infection of uninfected macrophages with extracellular *Legionella* may be derived in a probabilistic way. Assume that the number of contacts, $K$, that an extracellular *Legionella* has with other individuals (i.e., uninfected macrophages, macrophages infected with *Legionella* and extracellular *Legionella*) is Poisson distributed with mean $k$ contacts per unit time, $K \sim \text{Pois}(k)$. Given that the proportion of uninfected macrophages in the total system population is $U/N$, we assume a probability of $U/N$ that a *Legionella* contacts an uninfected macrophage once it comes to contact with another entity. The number of contacts, $n$, that a *Legionella* has with an uninfected macrophage during a time interval $(t, t + \delta t)$ follows a binomial distribution with probability $U/N$ and number of contacts following a Poisson distribution with mean $k\delta t$. This binomial-Poisson mixture results in a Poisson distribution with mean $kU\delta t/N$, calculated as follows:

$$\begin{aligned}
\mathbb{P}(n = i) &= \sum_{j=i}^{\infty} \mathbb{P}(n = i|K = j)\mathbb{P}(K = j), \\
&= \sum_{j=i}^{\infty} \binom{j}{i} \left(\frac{U}{N}\right)^{i} \left(1 - \frac{U}{N}\right)^{j-i} \frac{e^{-k\delta t}(k\delta t)^{j}}{j!}, \\
&= \left(\frac{U}{N}\right)^{i} \frac{(k\delta t)^{i}}{i! e^{k\delta t}} \sum_{j=i}^{\infty} \left(1 - \frac{U}{N}\right)^{j-i} \frac{(k\delta t)^{j-i}}{(j-i)!}, \\
&= \frac{(kU\delta t/N)^{i}}{i!} e^{-kU\delta t/N}, \\
&\implies n \sim \text{Pois}(kU\delta t/N).
\end{aligned}$$

Next, define $c$ as the probability of phagocytosis from a contact between an uninfected macrophage and an extracellular *Legionella*. We calculate the probability that phagocytosis of an extracellular *Legionella* occurs during any of its contacts as follows:

$$\begin{aligned}
\mathbb{P}(\text{Phagocytosised}) &= 1 - \mathbb{P}(\text{No contact causes phagocytosis}), \\
&= 1 - \sum_{i=0}^{\infty} (1 - c)^{i} \mathbb{P}(n = i), \\
&= 1 - \sum_{i=0}^{\infty} \frac{(1 - c)^{i}}{i!} \left(\frac{kU\delta t}{N}\right)^{i} e^{\frac{-kU\delta t}{N}}, \\
&= 1 - e^{\frac{-kU\delta t}{N}} \sum_{i=0}^{\infty} \frac{((1 - c)kU\delta t/N)^{i}}{i!}, \\
&= 1 - e^{-\frac{ckU\delta t}{N}}.
\end{aligned}$$

Further, by taking the Taylor series expansion for $e$, dividing by $\delta t$ and taking the limit as $\delta t \to 0$, a rate of infection for a single uninfected macrophage is obtained:

$$\text{Rate of Phagocytosis} = \frac{ckU}{N} \implies \frac{dU}{dt} = -\frac{ckLU}{N}.$$

Before continuing, we consider extending the assumption of a Poisson distribution for the number of contacts to allow for variability in $k$, in order to assess the effect that this variability has on $n$. We model the Poisson mean as a gamma-distributed random variable (or exponentially distributed, with a gamma distribution shape parameter equal to one). Therefore, we assume $K \sim \text{Gamma}(r, p)$. We define $Y = kU\delta t/N$ where $a = \frac{U\delta t}{N}$. Further, we consider an arbitrary point $y = a\hat{k}$:

$$\begin{aligned} F_Y(y) &= P(Y \le y) = P(\hat{k} \le y/a) = F_k(y/a), \\ \frac{dF_k(y/a)}{dy} &= \frac{dk}{dy}\frac{dF_k(k)}{dk} = \frac{(p/a)^r}{\Gamma(r)} y^{r-1} e^{-(p/a)y}, \\ &\implies Y \sim \text{Gamma}(r, q = p/a). \end{aligned}$$

From this, we calculate $n$ as a mixture $N|K \sim \text{Pois}(k)$ with $K \sim \text{Gamma}(r, q = p/a)$:

$$\begin{aligned} \mathbb{P}(N = i) &= \int_0^\infty \mathbb{P}(N|Y = y)\mathbb{P}(Y = y)dy, \\ &= \int_0^\infty \frac{y^i}{i!} e^{-y} \frac{(p/a)^r}{\Gamma(r)} y^{r-1} e^{-py/a} dy, \\ &= \frac{(p/a)^r}{i!\Gamma(r)} \int_0^\infty y^{i+r-1} e^{-(1+p/a)y} dy, \\ &= \frac{(p/a)^r}{i!\Gamma(r)} \frac{\Gamma(i+r)}{(1+p/a)^{i+r}}, \\ &= \frac{\Gamma(i+r)}{i!\Gamma(r)} \left(\frac{p/a}{1+p/a}\right)^r \left(\frac{1}{1+p/a}\right)^i, \\ &= \binom{i+r-1}{i} \left(\frac{p}{p+a}\right)^r \left(\frac{a}{p+a}\right)^i. \end{aligned}$$

Therefore, $n \sim \text{NB}(r, \frac{p}{p+a})$. The probability of phagocytosis occurring is calculated:

$$\begin{aligned} \mathbb{P}(\text{Phagocytosised}) &= 1 - \sum_{i=0}^{\infty} (1-c)^i \binom{i+r-1}{i} \left(\frac{q}{q+1}\right)^r \left(\frac{1}{q+1}\right)^i, \\ &= 1 - \left(\frac{q}{q+1}\right)^r \sum_{i=0}^{\infty} \left(\frac{1-c}{q+1}\right)^i \binom{i+r-1}{i}, \\ &= 1 - \left(\frac{q}{q+1}\right)^r \left[\left(1 - \frac{1-c}{q+1}\right)^{-r}\right], \\ &= 1 - \left(\frac{q}{q+c}\right)^r, \\ &= 1 - \left(\frac{pN}{pN + Uc\delta t}\right)^r, \\ &= 1 - \left[\sum_{i=0}^{\infty} \left(-\frac{Uc\delta t}{pN}\right)^i\right]^r \approx \frac{Urc\delta t}{pN}. \end{aligned}$$

Taking the limit at $\delta t \to 0$ leads to a rate of phagocytosis for an individual extracellular *Legionella* bacteria proportional to $U/N$. Accounting for variability in the number of contacts by allowing $k$ to be gamma distributed results in a probability which is equal to $U/N$ multiplied by a combination of the parameters. This extension can be considered the same as the original model where $k$ did not vary as the rate of phagocytosis derived for the deterministic system of ODEs is a superparameter (based on a multiple combination of these parameter) multiplied by $UL/N$. Hence, when deriving a system of ordinary differential equations both models are equivalent.

##### System of Equations

A deterministic system of ordinary differential equations may now be obtained to model the populations of both the uninfected and infected macrophages as well as the extracellular *Legionella*. First, we define $\epsilon$ as the growth rate of uninfected macrophages in the lungs. Second, we define $\beta$ as the rate of phagocytosis of extracellular *Legionella* in which the *Legionella* are killed. Third, we define $\alpha$ as the rate of phagocytosis of extracellular *Legionella* in which the *Legionella* survives and lives within the macrophage. Fourth, we define $\lambda$ as the rate at which an infected macrophage containing *Legionella* rupture and release *Legionella* back within the extracellular region of the lungs. Fifth, we define $G$ as the average number of *Legionella* released during the rupture event:

$$\begin{cases} \frac{dU}{dt} = \epsilon U - \frac{\alpha LU}{U+M+L}, \\ \frac{dM}{dt} = \frac{\alpha LU}{U+M+L} - \lambda M, \\ \frac{dL}{dt} = \lambda GM - \frac{\gamma LU}{U+M+L}. \end{cases} \tag{1}$$

This system of nonlinear equations can not be solved for an analytical solution. However, by considering a scenario in which a large uninfected macrophage population exists, (1) reduces to a linear system that may be solved with initial conditions $M(0) = M_0$ and $L(0) = L_0$:

$$\begin{cases} \frac{dM}{dt} = \alpha L - \lambda M, \\ \frac{dL}{dt} = \lambda GM - (\alpha + \beta)L. \end{cases} \tag{2}$$

Solving this system by considering eigenvalues and eigenvectors can be done as follows. Consider system (2) in matrix form:

$$\begin{pmatrix} \dot{M} \\ \dot{L} \end{pmatrix} = A \begin{pmatrix} M \\ L \end{pmatrix}, \quad \text{where} \quad A = \begin{pmatrix} -\lambda & \alpha \\ \lambda G & -\gamma \end{pmatrix}. \tag{3}$$

Eigenvalues of this system are calculated from solving the characteristic equation on this $2 \times 2$ matrix:

$$\begin{aligned} \det(xI - A) = 0 &\implies \det \begin{pmatrix} x+\lambda & -\alpha \\ -\lambda G & x+\gamma \end{pmatrix} = 0, \\ &\implies x^2 + (\lambda + \gamma)x + \lambda\gamma - \lambda\alpha G = 0, \\ &\implies x = \frac{1}{2}\left(-\lambda - \gamma \pm \sqrt{(\lambda - \gamma)^2 + 4\lambda\alpha G}\right). \end{aligned} \tag{4}$$

The corresponding eigenvectors of this system will be calculated. For simplicity of notation define $\theta = x_1 - x_2 = \sqrt{(\lambda - \gamma)^2 + 4\lambda\alpha G}$, so that $x_{1,2} = \frac{1}{2}(-\lambda - \gamma \pm \theta)$:

$$\text{(1)} \qquad \begin{pmatrix} -\lambda & \alpha \\ \lambda G & -\gamma \end{pmatrix} \begin{pmatrix} z_1 \\ z_2 \end{pmatrix} = \begin{pmatrix} x_1 z_1 \\ x_1 z_2 \end{pmatrix} \implies \begin{cases} (\lambda + x_1)z_1 = \alpha z_2, \\ (x_1 + \gamma)z_2 = \lambda G z_1, \end{cases}$$

$$\implies \frac{\alpha}{\lambda + x_1} = \frac{x_1 + \gamma}{\lambda G}. \tag{5}$$

$$\text{(2)} \qquad \begin{pmatrix} -\lambda & \alpha \\ \lambda G & -\gamma \end{pmatrix} \begin{pmatrix} z_1 \\ z_2 \end{pmatrix} = \begin{pmatrix} x_2 z_1 \\ x_2 z_2 \end{pmatrix} \implies \begin{cases} (\lambda + x_2)z_1 = \alpha z_2, \\ (x_2 + \gamma)z_2 = \lambda G z_1, \end{cases}$$

$$\implies \frac{\alpha}{\lambda + x_2} = \frac{x_2 + \gamma}{\lambda G}. \tag{6}$$

Therefore, the eigenvectors corresponding to $x_1$ and $x_2$ respectively are $\left(1, \frac{\lambda + x_1}{\alpha}\right)^T$ and $\left(1, \frac{\lambda + x_2}{\alpha}\right)^T$. Hence, this system can be solved with constants $C_1$ and $C_2$, in which these constants are calculated using the initial conditions:

$$\begin{pmatrix} P \\ L \end{pmatrix} = C_1 e^{x_1 t} \begin{pmatrix} 1 \\ \frac{\lambda + x_1}{\alpha} \end{pmatrix} + C_2 e^{x_2 t} \begin{pmatrix} 1 \\ \frac{\lambda + x_2}{\alpha} \end{pmatrix}.$$

Setting $t = 0$ gives $M_0 = C_1 + C_2$ and $L_0 = \frac{1}{\alpha}(C_1(\lambda + x_1) + C_2(\lambda + x_2))$. Solutions of these simultaneous equations give the system of equations defined as follows:

$$\begin{cases} M(t) &= \frac{\alpha L_0 - M_0(\lambda + x_2)}{\theta} e^{x_1 t} + \frac{M_0(\lambda + x_1) - \alpha L_0}{\theta} e^{x_2 t}, \\ L(t) &= \frac{\alpha L_0 - M_0(\lambda + x_2)}{\theta\alpha}(\lambda + x_1) e^{x_1 t} + \frac{M_0(\lambda + x_1) - \alpha L_0}{\theta\alpha}(\lambda + x_2) e^{x_2 t}. \end{cases} \tag{7}$$

#### Parameter dependency map

Due to the lack of experimental data, we could not de-couple all aspects of the within-host model and estimate each parameter independently of the others. Therefore, we introduce a parameter dependency map, as some of the parameter estimation process depends on previously estimated parameters. Here, we provide a figure that shows how the parameters of the within-host models depend on each other (Fig. 1)

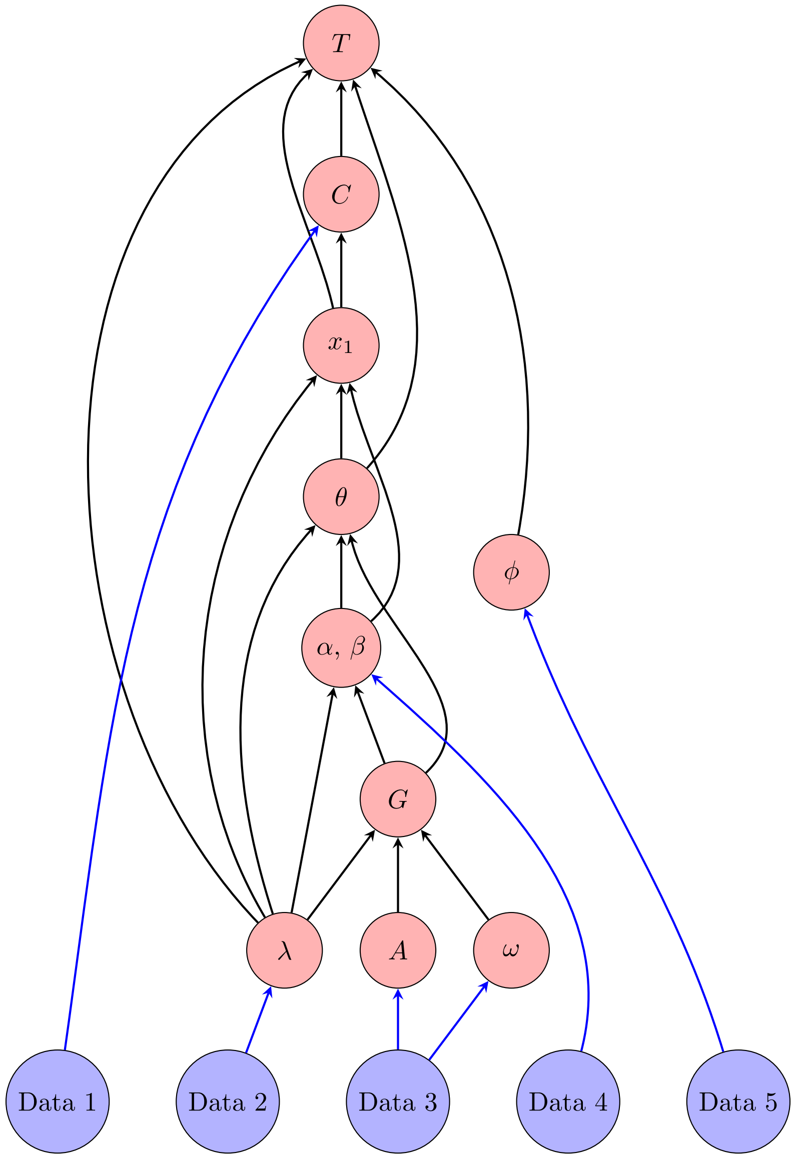


Figure 1: A parameter-dependency plot. In this work, we use five datasets for the within-host model parameterisation. Blue arrows indicate that the given dataset (located at the source of arrow) is used to estimate the given parameter (located at the head of the arrow). Additionally, black arrows from one parameter to another indicates that the parameter located at the head of the arrow depends on the estimate of the parameter located at the source of the arrow.

#### Parameter kernel plots

During the parameterisation process of the within-host parameters we generated distributions for each model parameter based on bootstrapping approaches, assumptions of normality on parameter estimates of nonlinear least square models and following the parameter dependencies provided above (Fig. 2).

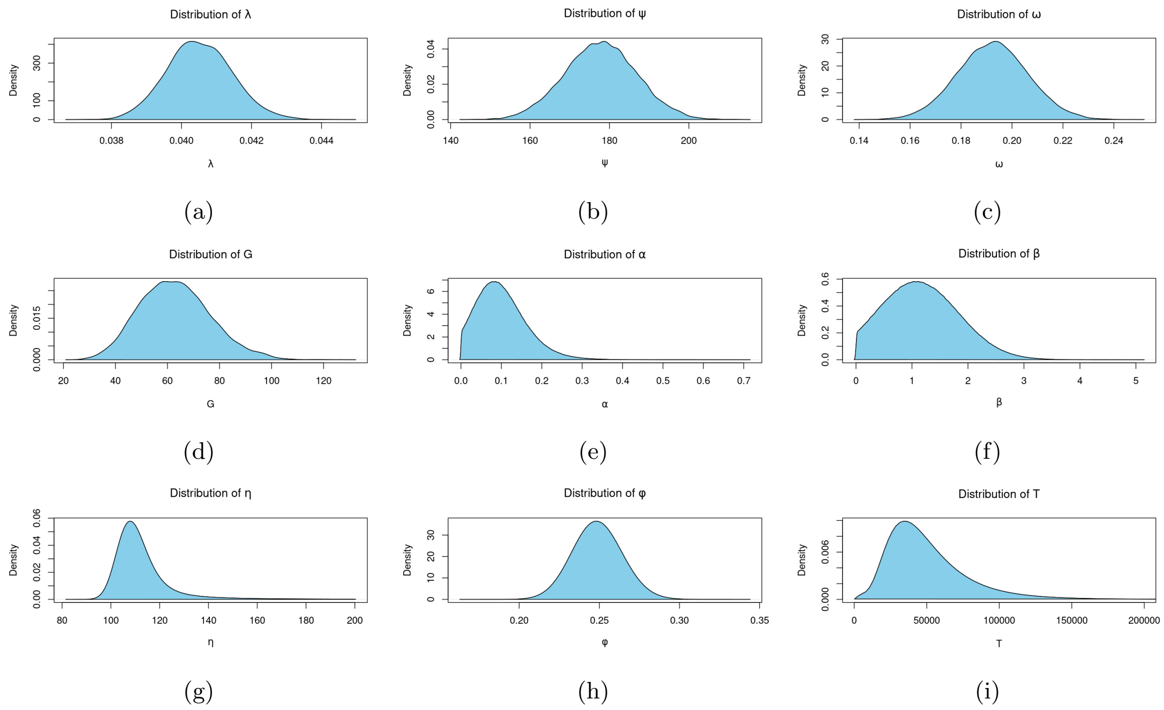


Figure 2: Kernel density plots that estimate the distribution for each parameter used in the within-host C model that models heterogeneity at the cellular and population levels.

#### Standard errors for parameter estimates obtained in DR and TDR analysis

Here, we provide the standard errors for the parameter estimates that were obtained when fitting the DR and TDR models the the simulated data within the paper. We begin with the DR pararmeter standard errors in Table 1.

| DR model | Stochastic Model | | | | | |
|---|---|---|---|---|---|---|
| | A | | B | | C | |
| | Standard error | Parameter estimates | Standard error | Parameter estimates | Standard error | Parameter estimates |
| Exponential | $d_{50} = 8.84$ | (0.02) | $d_{50} = 8.95$ | (0.02) | $d_{50} = 8.97$ | (0.02) |
| Beta-Poisson | $\alpha_b = 212.62$ | (227.7) | $\alpha_b = 1190$ | (6137) | $\alpha_b = 17.61$ | (1.51) |
| | $\beta_b = 2489.22$ | (2674.7) | $\beta_b = 14166$ | (73086) | $\beta_b = 201.42$ | (17.97) |
| Hill | $\alpha_h = 1.76$ | (0.02) | $\alpha_h = 1.77$ | (0.02) | $\alpha_h = 1.72$ | (0.02) |
| | $d_{50} = 8.54$ | (0.08) | $d_{50} = 8.66$ | (0.07) | $d_{50} = 8.52$ | (0.08) |
| Burr 1 | $\alpha_B = 0.79$ | (0.01) | $\alpha_B = 0.87$ | (0.01) | $\alpha_B = 0.81$ | (0.01) |
| | $\beta_B = 0.06$ | (0.001) | $\beta_B = 0.06$ | (0.001) | $\beta_B = 0.06$ | (0.0001) |
| | $d_{50} = 8.90$ | (0.02) | $d_{50} = 9.01$ | (0.02) | $d_{50} = 8.84$ | (0.02) |
| Burr 2 | $\alpha_D = -0.14$ | (0.02) | $\alpha_D = 0.04$ | (0.03) | $\alpha_D = -0.19$ | (0.01) |
| | $\beta_D = 0.08$ | (0.002) | $\beta_D = 0.08$ | (0.001) | $\beta_D = 0.06$ | (0.06) |
| | $\gamma_D = -0.07$ | (0.01) | $\gamma_D = -0.13$ | (0.01) | $\gamma_D = -0.0001$ | (0.11) |
| | $d_{50} = 8.861$ | (0.03) | $d_{50} = 8.89$ | (0.03) | $d_{50} = 8.84$ | (0.02) |

Table 1: Results of fitting the DR models to the simulated data, with the parameter estimates and corresponding standard errors provided.

Similarly, we provide the parameter estimates and corresponding standard errors from fitting the TDR models to the simulated dose-dependent incubation-period data.

#### Moments analysis of dose-dependent incubation-period simulations

We provide analysis of the first four moments of the dose-dependent incubation-period data simulated from the three within-host models (Fig. 3).

| | Stochastic Model | | | | | |
|---|---|---|---|---|---|---|
| | Model A | | Model B | | Model C | |
| TDR model | Parameter estimates | Standard error | Parameter estimates | Standard error | Parameter estimates | Standard error |
| Exp w/ RT | $k_0 = 380.11$ | (0.322) | $k_0 = 367.55$ | (0.31) | $k_0 = 314.04$ | (0.13) |
| | $k_1 = 0.73$ | (0.005) | $k_1 = 0.61$ | (0.005) | $k_1 = -0.42$ | (0.002) |
| Exp w/ PT | $k_0 = 1.21 \times 10^6$ | $(1.88 \times 10^4)$ | $k_0 = -1.23 \times 10^6$ | $(1.87 \times 10^4)$ | $k_0 = 548.40$ | (4.15) |
| | $k_1 = -2.45$ | (0.002) | $k_1 = -2.48$ | (0.002) | $k_1 = -0.95$ | (0.006) |
| | $k_2 = 3.15$ | (0.004) | $k_2 = 3.17$ | (0.004) | $k_2 = 1.16$ | (0.002) |
| Exp w/ LT | $k_0 = 0.12$ | (0.0002) | $k_0 = 0.12$ | $(2.24 \times 10^{-4})$ | $k_0 = 0.07$ | $(7.70 \times 10^{-5})$ |
| | $\tau = 106.14$ | (0.08) | $\tau = 105.67$ | (0.08) | $\tau = 140.73$ | (0.08) |
| BP w/ RT | $\alpha = 80608.94$ | $(6.14 \times 10^4)$ | $\alpha = 40361.48$ | $(5.98 \times 10^4)$ | $\alpha = 2.83$ | (0.01) |
| | $j_0 = 380.11$ | (0.32) | $j_0 = 367.56$ | (0.31) | $j_0 = 328.64$ | (0.14) |
| | $j_1 = -1.10$ | (0.005) | $j_1 = -0.98$ | (0.005) | $j_1 = -0.25$ | (0.002) |
| BP w/ PT | $\alpha = 6.82 \times 10^4$ | $(3.97 \times 10^4)$ | $\alpha = 7.36 \times 10^4$ | $(3.84 \times 10^4)$ | $\alpha = 2.68$ | (0.01) |
| | $j_0 = 1.48 \times 10^6$ | $(2.34 \times 10^4)$ | $j_0 = 1.23 \times 10^6$ | $(1.86 \times 10^4)$ | $j_0 = 773.39$ | (5.52) |
| | $j_1 = 2.10$ | (0.002) | $j_1 = 2.12$ | (0.002) | $j_1 = 0.54$ | (0.005) |
| | $j_2 = 3.21$ | (0.004) | $j_2 = 3.17$ | (0.004) | $j_2 = 1.25$ | (0.002) |
| BP w/ LT | $\alpha = 6.71 \times 10^4$ | $(7.18 \times 10^4)$ | $\alpha = 9.08 \times 10^4$ | $(7.29 \times 10^4)$ | $\alpha = 2.42$ | (0.03) |
| | $j_0 = 0.12$ | $(2.24 \times 10^{-4})$ | $j_0 = 0.12$ | $(2.24 \times 10^{-4})$ | $j_0 = 0.08$ | $(1.13 \times 10^{-4}$ |
| | $j_1 = 12.86$ | (0.01) | $j_1 = 12.70$ | (0.01) | $j_1 = 10.24$ | (0.007) |
| Burr | $\alpha_D = -0.09$ | (0.007) | $\alpha_D = 0.91$ | (0.008) | $\alpha_D = -0.12$ | (0.003) |
| | $\beta_D = 0.03$ | (0.02) | $\beta_D = -0.71$ | (0.03) | $\beta_D = 0.04$ | $(1.83 \times 10^{-4})$ |
| | $\gamma_D = 0.002$ | (0.04) | $\gamma_D = 0.75$ | (0.03) | $\gamma_D = 1.42 \times 10^{-5}$ | $(2.16 \times 10^{-4})$ |
| | $d_{50} = 9.70$ | (0.01) | $d_{50} = 9.66$ | (0.02) | $d_{50} = 9.13$ | (0.01) |
| | $\alpha_I = 20.73$ | (0.15) | $\alpha_I = 19.41$ | (0.15) | $\alpha_I = 3.50$ | (0.01) |
| | $\beta_I = 264.77$ | $(1.779 \times 10^2)$ | $\beta_I = 58.19$ | (8.47) | $\beta_I = 18.15$ | (0.06) |
| | $\tau_1 = 207.17$ | (2.31) | $\tau_1 = 130.61$ | (0.35) | $\tau_1 = 142.54$ | (0.31) |
| | $\tau_2 = 0.06$ | $(8.83 \times 10^{-4})$ | $\tau_2 = 0.13$ | $(8.62 \times 10^{-4})$ | $\tau_2 = 0.13$ | $(6.90 \times 10^{-4})$ |
| | $\tau_3 = -93.79$ | (2.42) | $\tau_3 = -7.86$ | (0.48) | $\tau_3 = -12.90$ | (0.42) |

Table 2: The results of fitting both the data–driven and mechanistic TDR models to the simulated data, parameter estimates and corresponding standard errors provided.

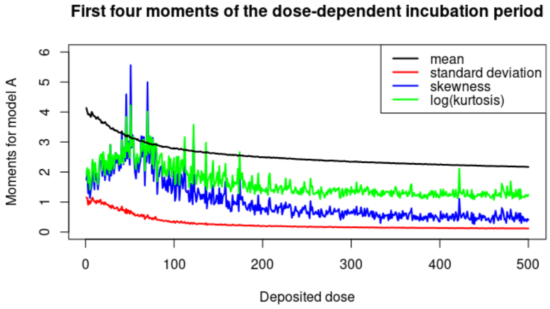


(a)

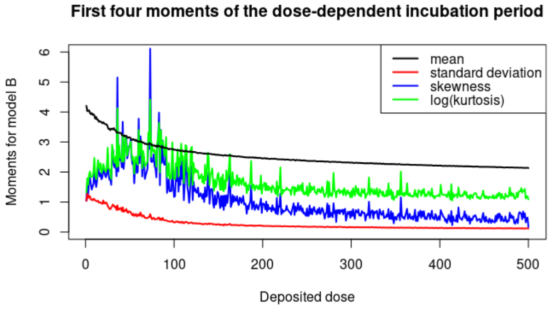


(b)

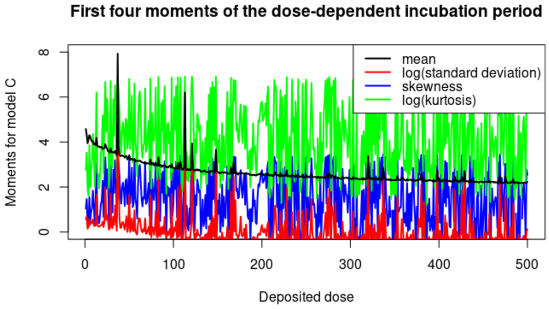


(c)

Figure 3: First four moments of the incubation period data generated from each of the within-host models.

For within-host models A and B the results are similar; the mean incubation period decreases as the deposited dose increases. For both within-host models, the kurtosis tends towards three for large deposited dose, indicating that the incubation period of Legionnaires' disease may be well approximated by a normal distribution for large deposited doses. However, to follow a normal distribution, the skewness of the data must tend to zero, which does not appear to be the case (Figs. 2(a), 2(b)). Furthermore, the within-host model C offers a larger variation in the results due to the assumption of heterogeneity in the population. The results of the within-host simulation largely depend on the values of the parameters sampled, which results in the observed variable moments across the deposited dose range (Fig. 2(c)). These results indicate that the deposited dose is less important for the heterogeneous model compared to the homogeneous models (within-host models A and B).

#### Hierarchical analysis

With the incubation-period data simulated from the three within-host models, we analyse how the deposited dose affects the incubation period. We use a hierarchical clustering analysis to group the deposited doses (i.e., low dose, medium-dose and high-dose incubation-period distributions) by comparing the incubation-period distributions for each deposited dose (Fig. 4).

We calculate the within-cluster sum of squares for a specified number of clusters $k$, and use Ward's linkage method for joining clusters, which focuses on minimizing the variance within clusters. For each number of cluster $k$ and within-cluster sum of square calculated we estimate the optimal $k$ using an elbow method from visualising the results plotted (Fig 4). For within-host model A, we obtain an optimal $k = 3$, which corresponds to three clusters (low-dose cluster, medium-dose cluster and high-dose cluster). Further, for within-host models B and C, we obtain an optimal $k = 4$, which corresponds to four clusters (low-dose cluster, low-medium-dose cluster, high-medium-dose cluster and high-dose cluster).

We test whether the clustered datasets may be interpreted as being produced from different distributions. Therefore, for each within-host model, we perform the Mann–Whitney U test on the clustered datasets. For within-host model A, we record a p-value of less than $10^{-15}$ when performing the test on the low-dose and medium-dose datasets, indicating that these two datasets are statistically significantly different. Additionally, a p-value that is similarly small (less than $10^{-15}$) when performing the test on the medium-dose and high-dose datasets. For the within-host model B, we record a p-value of less than $10^{-15}$ when conducting the Mann–Whitney U test on all possible pairs of datasets: low-dose with medium–low dose, medium–low dose with medium–high dose and medium–high dose with high dose. Similarly, for the within-host model C, we record a p-value of less than $10^{-15}$ when performing the Mann–Whitney U test on all possible pairs of datasets: low-dose with medium–low dose, medium–low dose with medium–high dose and medium–high dose with high

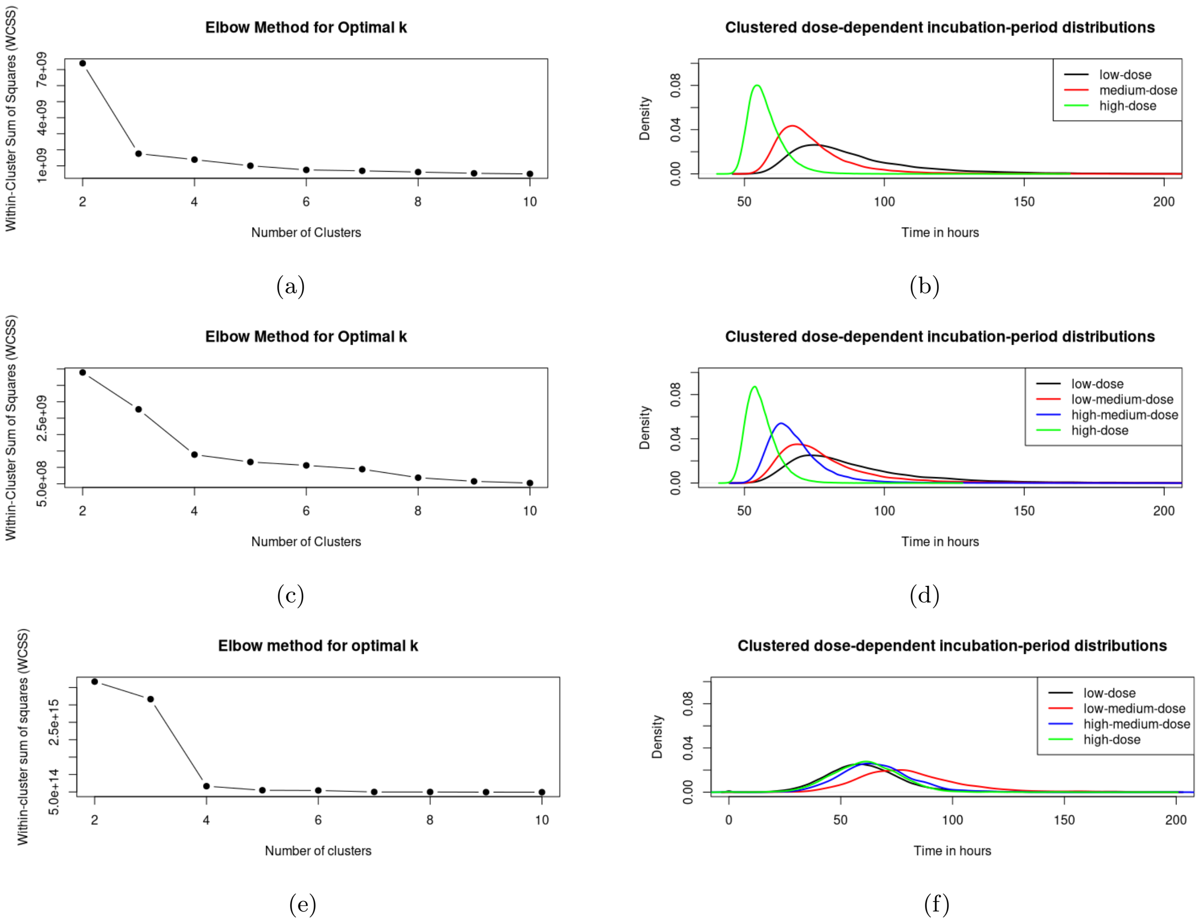


Figure 4: Elbow-finding procedure to find the optimal number of clusters for categorising the deposited doses based on simulated incubation periods. The method for finding the optimal number of clusters are provided in sub-figures (a), (c) and (e) for within-host models A, B and C respectively. Additionally, the clustered incubation-period distributions are provided in sub-figures (b), (d) and (f) for within-host models A, B and C respectively.

dose. Therefore, for all within-host models, all clustered incubation-period datasets are statistically significantly different from one another and we can group values of deposited doses to describe what kind of incubation period an individual can expect to endure.

#### Sensitivity analysis

To conduct the sensitivity analysis, we simulate the within-host model with a deposited dose equal to nine, as it is close to the ID50 estimated from all of the within-host models. The within-host model is simulated for each model parameter, which we sample from their respective distribution. For the chosen parameter, we fix all parameters that do not depend on the parameter chosen. Following this, we calculate point estimates for the parameters that depend on the chosen parameter to obtain estimates for the within-host model simulation, so that we are not introducing bias into our analysis. For each sample of our chosen parameter, we take 1000 simulations of the within-host model to record the proportion of occurrences in which symptom onset occurred (to be used as a proxy for probability of symptom onset) and the incubation period of each simulation that resulted in symptom onset. This process is repeated for one hundred samples of each parameter. With this generated data, we calculate Spearman's correlation to assess the strength of the monotonic relationship between the value of each variable in the within-host model and the simulation outputs (i.e, probability of symptom onset and incubation period), with results provided in Fig. 5.

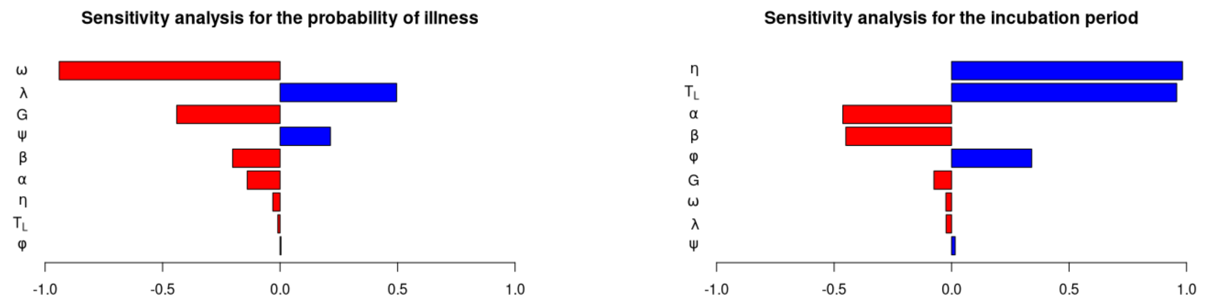


(a) (b)

Figure 5: Plots of a sensitivity analysis conducted for the within-host model.

In the analysis of probability of illness, the intracellular growth rate $\omega$ provided the strongest negative influence, while the infected-macrophage rupture rate $\lambda$ provided the strongest positive influence. As $\omega$ and $\psi$ were correlated, a smaller estimate for $\omega$ corresponded to a larger $\psi$, which meant that more intracellular *Legionella* may occupy an infected macrophage at full (or close to full) capacity. Furthermore, a faster rupture rate indicates that intracellular *Legionella* become free to occupy the extracellular region of the lungs faster after infection of a macrophage, resulting in larger extracellular populations earlier. This change means that the macrophage population may struggle to eliminate the total extracellular *Legionella* population, resulting in a larger probability of illness. The parameters $\eta$, $T_L$ and $\phi$ either directly or indirectly control the maximum inflammation levels within the lungs before onset of symptoms. Before an individual becomes symptomatic, they must reach a point of no return in which the extracellular *Legionella* population is sufficiently large that the process is approximately deterministic and illness of eventually inevitable. This point will occur much earlier than the maximum inflammation levels are reached. Therefore, $\eta$, $T_L$ and $\phi$ have little effect on the probability of illness. A surprising result is the negative correlation between $G$ and the probability of illness. If $G$ increases, this may alter the ratio $\alpha/(\alpha + \beta)$, which in turn affects the probability that illness occurs. Additionally, increasing both $\alpha$ and $\beta$ result in lower probability of illness, indicating that faster rate of phagocytosis is linked to a higher chance of bacterial clearance.

Next, for the analysis of the incubation period, $\eta$ and $T_L$ are directly related to the time until symptom onset, thus a strong positive correlation is expected. Additionally, a negative correlation between both $\alpha$ and $\beta$ with the incubation period is expected. A faster rate of phagocytosis means that events are occurring faster, and the population will reach $T_L$ earlier. Moreover, $\psi$, $\omega$, $\lambda$ and $G$ have a small correlation with the incubation period. Varying these values result in a different estimate of $T_L$, which indicate that they do not themselves are not important for the incubation period. Finally, $\phi$ is the probability of deposition and since we are modelling $t = 0$ as the scenario once *Legionella* are deposited, we expect a small correlation between $\phi$ and the incubation period.
